# Relationship between sex biases in gene expression and sex biases in autism and Alzheimer’s disease

**DOI:** 10.1101/2023.08.29.23294773

**Authors:** Stuart B. Fass, Bernard Mulvey, Wei Yang, Din Selmanovic, Sneha Chaturvedi, Eric Tycksen, Lauren A. Weiss, Joseph D. Dougherty

## Abstract

Sex differences in the brain may play an important role in sex-differential prevalence of neuropsychiatric conditions. In order to understand the transcriptional basis of sex differences, we analyzed multiple, large-scale, human postmortem brain RNA-seq datasets using both within-region and pan-regional frameworks. We find evidence of sex-biased transcription in many autosomal genes, some of which provide evidence for pathways and cell population differences between chromosomally male and female individuals. These analyses also highlight regional differences in the extent of sex-differential gene expression. We observe an increase in specific neuronal transcripts in male brains and an increase in immune and glial function-related transcripts in female brains. Integration with single-cell data suggests this corresponds to sex differences in cellular states rather than cell abundance. Integration with case-control gene expression studies suggests a female molecular predisposition towards Alzheimer’s disease, a female-biased disease. Autism, a male-biased diagnosis, does not exhibit a male predisposition pattern in our analysis. Finally, we provide region specific analyses of sex differences in brain gene expression to enable additional studies at the interface of gene expression and diagnostic differences.

**Graphical Abstract:** 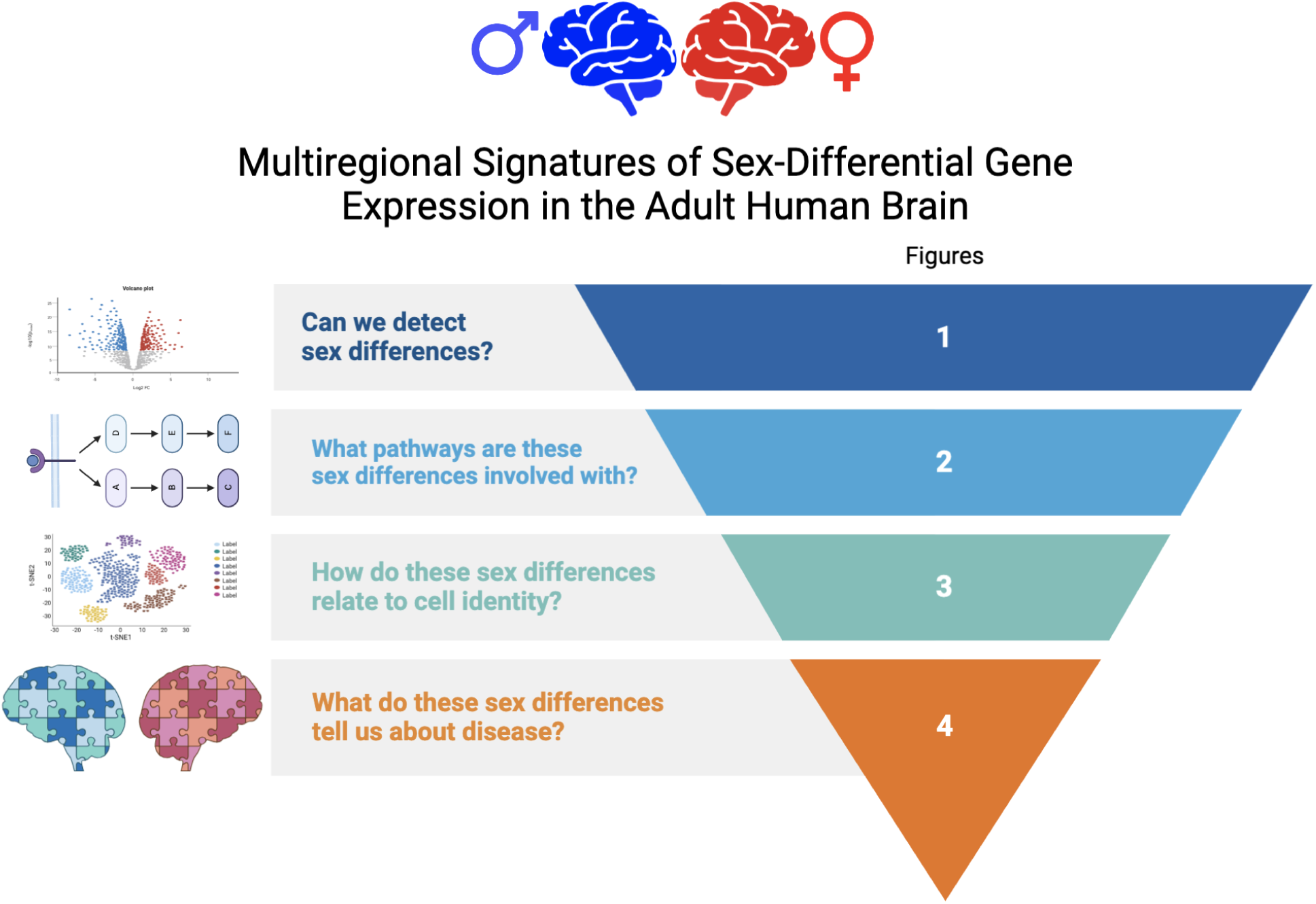

## Introduction

Most human neuropsychiatric conditions show differences in diagnostic rates between males and females. For example, males make up a higher percentage of diagnosed neurodevelopmental conditions that begin in early life, such as autism and Attention Deficit-Hyperactivity Disorder (ADHD). Conversely, females are more likely to suffer later-onset disorders such as Major Depressive Disorder and Alzheimer’s disease (Collaborators, GBD 2019 Diseases and Injuries et al., 2020; Collaborators, 2022). Furthermore, even though a major component of risk for these neuropsychiatric conditions is heritable, genetic risk is complex, with hundreds to thousands of genes and variants implicated (Consortium, 2017; Davies et al., 2017; Demontis et al., 2018; Howard et al., 2018; Consortium et al., 2019; Grove et al., 2019; Matoba et al., 2020; Ramaswami et al., 2020; Bellenguez et al., 2022; Mattheisen et al., 2022; Rajagopal et al., 2022). Moreover, depending on sex, manifestations of many disorders differ molecularly (Labonté et al., 2017; Seney et al., 2018; Kissel and Werling, 2020) and in their clinical presentation (Marcus et al., 2005; Arnett et al., 2015; Rubinow and Schmidt, 2018). Overall, the multifactorial heritability patterns and heterogenous phenotypes of neuropsychiatric conditions have been a substantial barrier toward understanding the biological processes governing sex differences in the odds of developing each condition and in their presentations.

Each of these sex-biased conditions is strongly influenced by large numbers of common, non-coding variants in the genome (Consortium, 2017; Demontis et al., 2018; Howard et al., 2018; Consortium et al., 2019; Grove et al., 2019; Matoba et al., 2020; Bellenguez et al., 2022). Common variants are thought to influence risk of psychiatric conditions by subtly affecting the expression of nearby genes in the brain. These many small changes in gene expression can, in aggregate, greatly alter risk of presenting with a given condition (O’Brien et al., 2018; Song et al., 2019; Dong et al., 2021), thus giving gene expression an important role in pathogenesis and progression.

Sex differences in transcription are modulated by several classes of DNA-interacting proteins, including those encoded on allosomes (X and Y chromosomes). Some DNA-interacting proteins are also modulated by sex-differential small molecule signals, including androgens, estrogens, and progestins. Beyond the direct binding sites of allosome-encoded proteins or sex hormone receptors, these primary factors appear to also have prominent roles in sex-differential transcriptional regulation via indirect and co-regulatory activity, including at risk variants for sex-biased psychiatric conditions (Bernabeu et al., 2021; Martin et al., 2021; Benjamin et al., 2022; Zhu et al., 2022; Mulvey et al., 2023). Ultimately, understanding the full extent of how and where sex governs transcription will improve the understanding of how gene expression affects the odds of developing a particular neuropsychiatric condition, the condition’s presentation, and how sex might relate to the expression differences due to genetic variants associated with each.

Autism is particularly illustrative of the complex interaction between sex and genetic risk. Researchers have proposed that subtle baseline sex differences in gene expression can shift the brain towards a transcriptomic signature that might promote the condition in a particular sex (Werling and Geschwind, 2013, 2013; Kissel and Werling, 2020) – *i.e.,* perhaps the male brain at baseline has a ‘molecular predisposition’ towards autism, and thus it takes fewer heritable genetic or environmental factors to meet criteria for a diagnosis, as posited by the “extreme male brain” theory (Baron-Cohen, 2002). If indeed case-control differences in transcription highlight such a state, a prediction of this model is that transcription in the male brain at baseline will be more similar to what is seen in postmortem autism brains. Indeed, past work found that upregulated genes in the neurotypical postmortem male (vs. female) cortex are more highly expressed in the postmortem cortex of autism patients from both sexes when compared to controls (Werling et al., 2016).

In addition to the effects of common variants on risk, rare loss of function mutations– implying a 50% reduction in expression– also cause disease, particularly for syndromic forms of autism and Intellectual Disability where hundreds of new causal genes have recently been identified (Abrahams et al., 2013; Fu et al., 2022). Thus, it would be interesting to examine whether any sex biases in gene expression overlap with rare-variant disorder genes. Previous work found that genes implicated in rare-variant forms of autism at that time did not show any sex bias in expression (Werling et al., 2016).

Therefore, to replicate and extend these prior studies we further characterized the transcriptomes of adult brains using larger datasets and additional brain regions, and tested whether sex-differential expression (DE) of risk genes themselves may underlie sex differences in incidence of two prominent sex-biased conditions - one male-biased (autism) and one female-biased (Alzheimer’s disease), both of which were selected because they have robust genome-wide association studies (GWAS) and case-control gene expression data. We examined two of the largest collections of postmortem brain RNA-seq data available: GTEx version 8 (Carithers et al., 2015) and the CommonMind Consortium (CMC) (Hoffman et al., 2019). A key advantage of the GTEx dataset is that it surveys multiple brain regions across hundreds of male and female individuals, enabling an analysis for sex both within and across brain regions. The CMC dataset consists of only frontocortical samples, which we used to benchmark our analysis of GTEx cortex and produce a high-confidence meta-analyzed set of sex-differentially expressed cortical genes. We thus present a resource grounded in a large bulk RNA-seq brain dataset, detailing sex-differential expression in the human adult brain at both broad and fine scale. Using signal-to-noise ratio (SNR) analyses, we identify regions with the most robust transcriptome-wide DE signatures within the GTEx data. We then identify differentially expressed genes (DEGs) in a novel ‘omnibus’ brain-wide framework, as well as DEGs for each region individually. With our omnibus analysis we identify a substantial proportion of the transcriptome as being sex-DE, albeit at very small magnitudes. From omnibus and regional sex DEGs, we then identify pathways and cell types over-or under-represented in each sex. We also integrate these results with insights from recent human single-cell RNA-seq data, which provide more refined cell type and subtype gene signatures. Finally, we examine whether baseline sex DE overlaps with rare and common variant disease-associated gene sets and DEGs from postmortem human brain studies of neuropsychiatric cohorts.

## METHODS

### GTEx

#### Data pre-processing, filtering, and normalization

Bulk RNA-sequencing data was previously conducted by GTEx and the Commonmind Consortium Except where noted, the GTEx data and CMC data were handled identically. We used GTEx project v8 release gene count data (annotated with Gencode v26). Sample and donor attribute files were downloaded (dbGaP: phs000424.v9.p2); count matrix and metadata tables for samples were passed to the R package *edgeR* (Robinson et al., 2009) to construct a *DGEList()* for downstream analyses. All donors coded either as having an unknown status or positive diagnosis for brain-related diseases were removed. These included ALS, Alzheimer’s disease, dementia, encephalitis at death, Creutzfeld-Jakob disease, multiple sclerosis, Parkinson’s disease, Reye’s syndrome, and schizophrenia. We additionally removed donors positive or of unknown status for major systemic diseases with potential to impact the brain secondarily in line with prior analyses (Hartl et al., 2021): sepsis/positive blood cultures, lupus, cardiovascular disease, HIV, active cancer diagnosis, high unexplained fever, abnormal white blood cells, influenza, and opportunistic infections. Overall this resulted in the sample numbers and sex distribution described in **Table 1**. We filtered out low-representation genes using *edgeR’s filterByExpr()* function, retaining those with greater than 10 counts per million (CPM) in 19 samples: 70% of the smallest group size of 27 (female amygdala). Data was weighted and scaled by library size with the TMM method using *edgeR’s calcNormFactors()* TMM method.

**Table 1:**
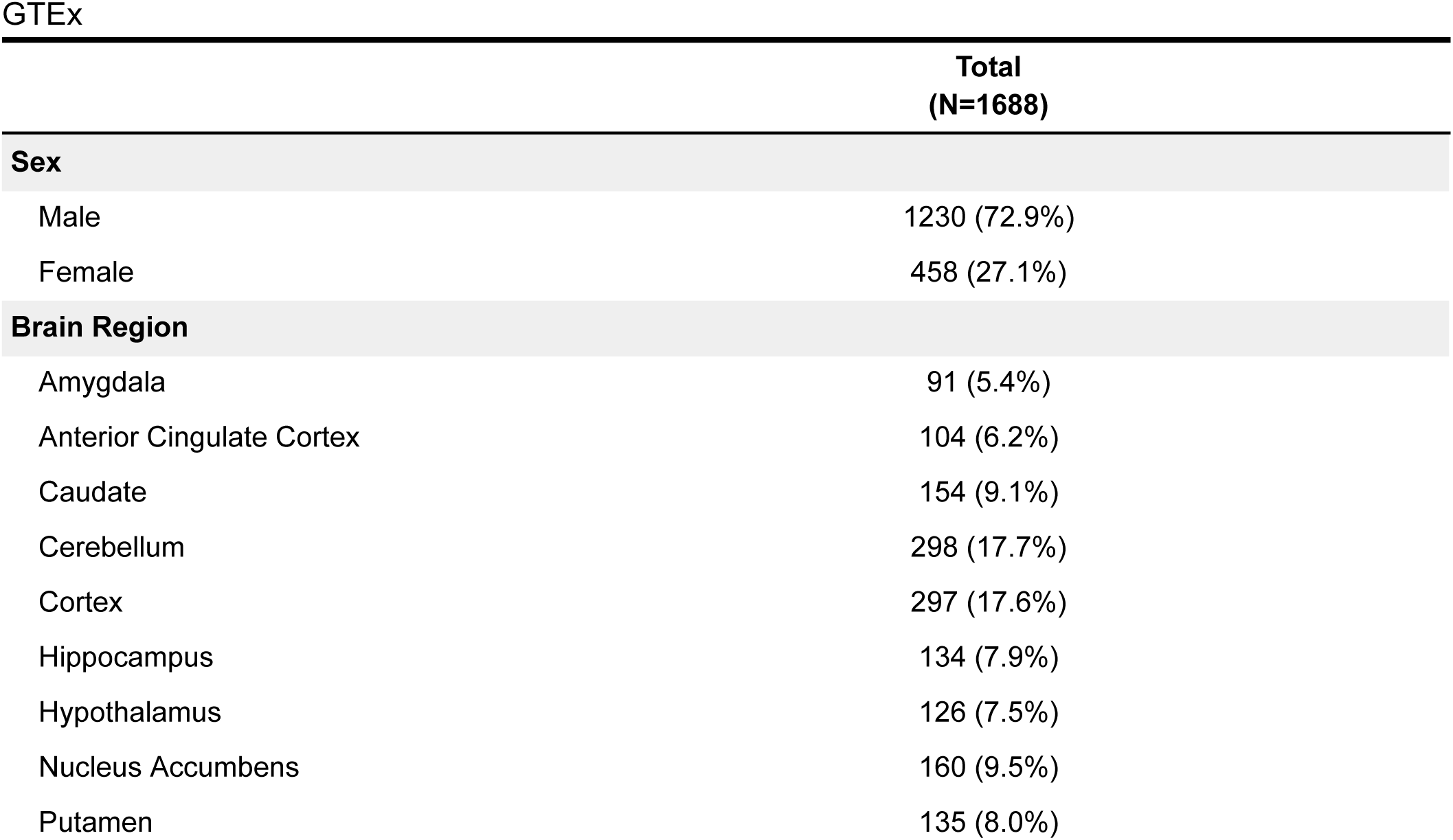
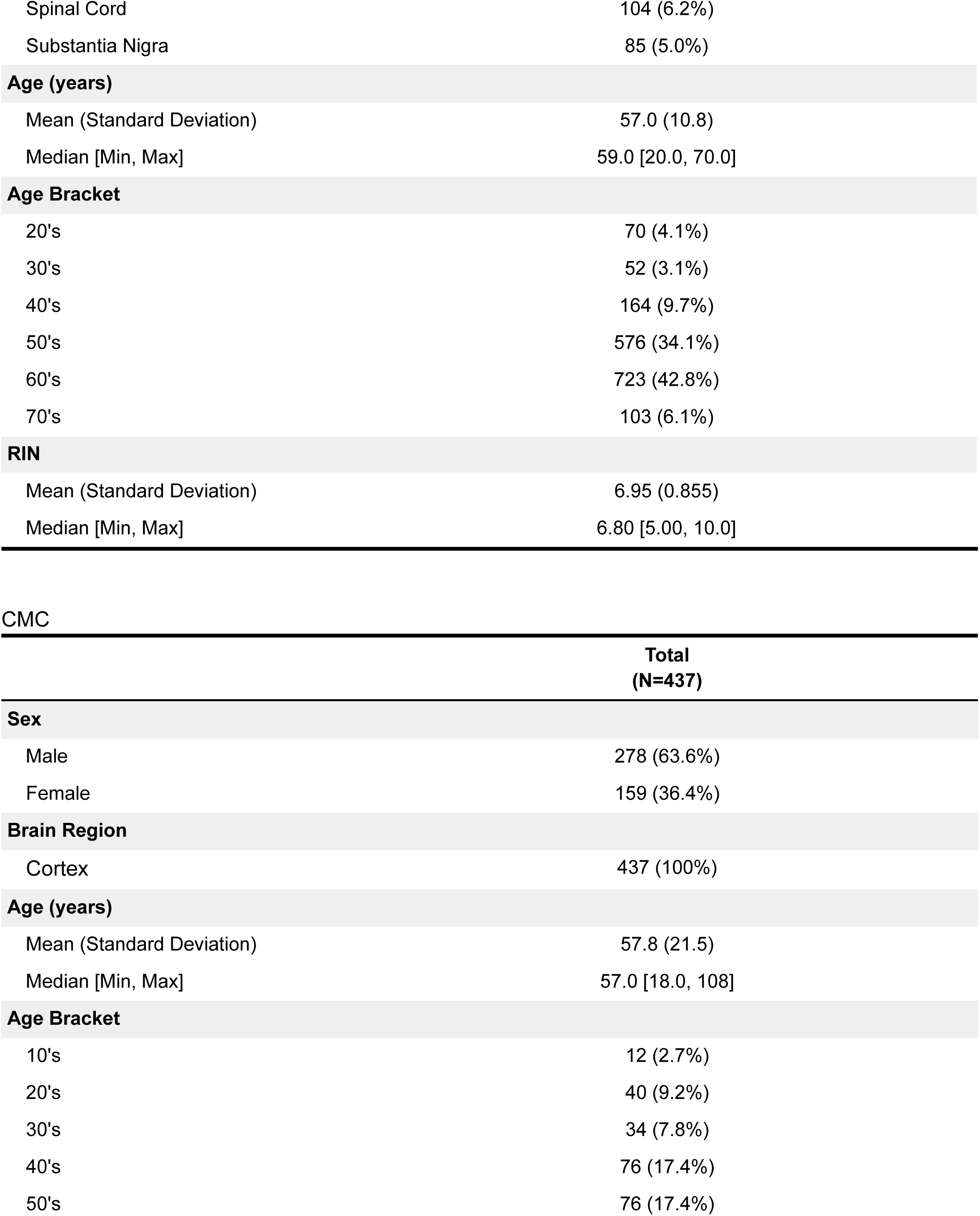
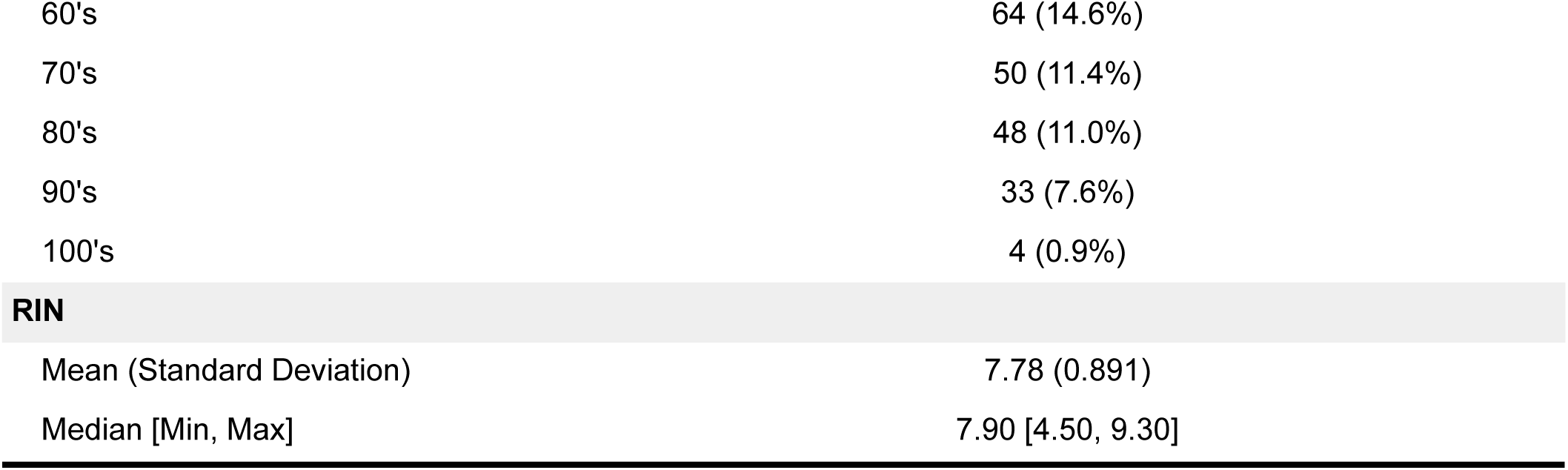
Donor demographic and sample quality information from GTEx and CMC datasets.

#### Surrogate Variable (SV) Analysis

Given the broad number of epidemiologic variables in the GTEx cohort, surrogate variables (SVs) were included to account for unknown latent sources of variation in the data. Forty nine SVs were identified using the svaseq() function from the R package *sva* (Leek and Storey, 2007). The number of SVs to utilize were calculated using the *num.sv()* function (“be” method). The full model and null model used for this function are as follows were:

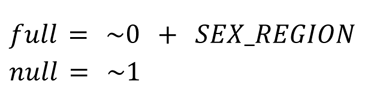

#### QC

PCA plots were generated using the *prcomp()* method. The effect of SVs was visualized using the *removeBatchEffect()* function from the *limma* R package (Liu et al., 2015; Ritchie et al., 2015). SVs were tested for correlation with any sex-region group in the data. Distributions of PCs generated from the counts matrix were plotted before and after SV correction to ensure SVs were not erroneously grouping our samples or creating outliers. Mean-variance trends were plotted and inspected visually to ensure genes were following typical trends for a sizable multigroup RNAseq experiment, namely ensuring there are no outlier genes relative to the mean-variance trend line, and that genewise dispersions were in the .5 to 1.5 range (**Supplemental Figure 1**).

#### Signal to noise ratio

Due to the high degree of variability inherent in obtaining postmortem brain tissue, it was critical that we were able to determine whether the influence of our biological signal of interest (sex) was detectable over technical noise. To evaluate this, we compared the total differences between males and females to the total variance in the unadjusted data according to the methods of Lopes-Ramos (Lopes-Ramos et al., 2020) to calculate a signal-to-noise ratio (SNR). Let F denote the number of females and M denote the number of males, and let X and Y be the matrices of gene expression values in females and males respectively, and let *X* and *Y* be the genewise expression average across all female and male samples. The exact equation for SNR calculations are as follows:

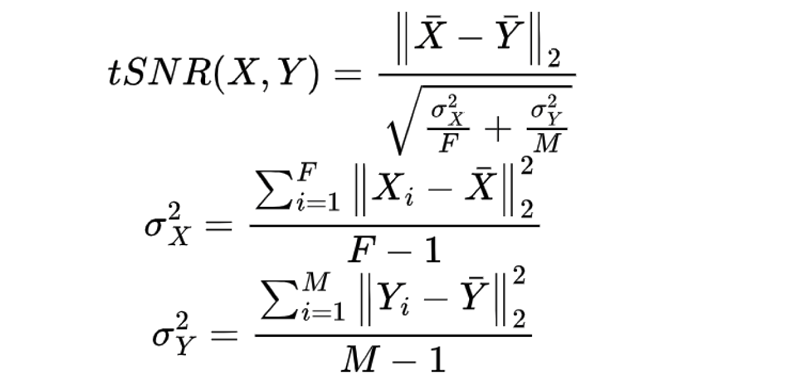

In order to determine SNR values without bias for unbalanced representation of sexes in the data, we calculated the SNR 10,000 times based on randomly drawn samples with correct sex labels to generate a “true” distribution, then calculated this value 10,000 more times from randomly drawn samples that were split into two arbitrary groups and randomly assigned a sex to create a null distribution. For each iteration per region, where n = 90% of the smaller (in all cases, female) group size, *n* male samples and *n* female samples were drawn (or for null simulations, 2n of the total samples). For a hypothetical brain region with 200 male samples and 100 female samples, 90 male and 90 female samples would be randomly selected per ‘true’ iteration and 180 samples selected and randomly assigned a sex per null iteration. We compared the distributions with Wilcox tests, and also calculated empirical P values as follows:

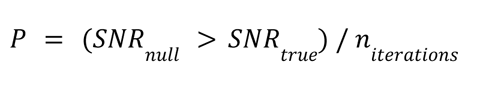

#### Differential expression analysis

Linear model designs were created using the base R *model.matrix()* function. The model used each sex-region (group) and the remaining SVs as fixed effects and donor as a random effect. Note that random effects (otherwise known as blocking factors) were estimated using a parallel implementation of the *limma* function *duplicateCorrelation()* (see *Code and Modules below*).

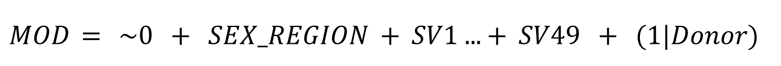

After the model was created, the count matrix was transformed to moderated log2 counts-per-million with a parallelized implementation *limma’s* the *VoomWithQualityWeights()* function so that run times were reduced. This function differs by utilizing the *arrayWeightsQuick()*, and a custom parallel *duplicateCorrelation()* function to be able to estimate random effects. As is the case with the packaged *VoomWithQualityWeights()* function, voom is run twice, once without the weights, followed by estimation of the sample weights, then voom is run again including the weights. While the packaged *VoomWithQualityWeights()* function then calculates row wise or REML-based gene weights, at this step we instead use another function from the *limma* package, *arrayWeightsQuick()*. Voom is run a second time with the updated weights, then the weights are estimated once more using the custom parallelized *duplicateCorrelation()*, followed again by *ArrayWeightsQuick()*. Finally, the counts matrix is adjusted based on these weights. These functions fit the counts matrix to a linear model estimating random factors, and adjust the relative weight of each sample relative to the mean variance of the sample. This means samples with high variance relative to the mean variance of all the samples will have less weight when detecting differentially expressed (DE) genes. This is a good replacement for trying to model the quality of the samples with RIN scores, which is generally a poor estimator (Jaffe et al., 2017). The linear “mixed” model is then fit to the adjusted data using the *lmfit()* in *limma*. Our dataset-wide modeling strategy allows for regional and omnibus contrasts using the same model, yielding regionally comparable results and precluding the need for multiple-testing correction for multiple contrasts. The following contrasts were used:

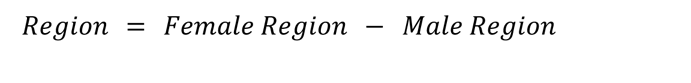

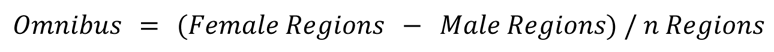

Therefore, all logFC values reported are positive for greater expression in females relative to males, and negative for greater expression in males vs females. The linear model-adjusted data are then contrasted to calculate differential expression and smoothed using *limma’*s *Ebayes()* function with default parameters, which utilizes an empirical Bayes method to squeeze the genewise-wise residual variances of the data towards a common value and provide a better estimate of the t-statistic than an unmoderated version. The DE tables for each region and the omnibus contrast are provided in **Supplemental Table 1**.

### CMC

#### Data wrangling, filtering, normalization

All wrangling and gene filtering done in the CMC dataset were performed in the same manner as for the GTEx dataset. Genes were annotated with GRCh37 using the tools from the Ensembl R package. All samples with a Schizophrenia or Klinefelter diagnosis were removed from the analysis. Overall this resulted in the sample numbers and sex distribution described in **Table 1**.

#### SVA

Again using the *sva* package’s svaseq function, we identified surrogate variables (SVs) representing latent sources of variation, all 9 of which were included in the DE model. The full model and null model provided to svaseq were as follows:

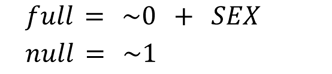

#### Differential expression analysis

Data was analyzed in the same manner as GTEx, except for use of the limma’s packaged VoomWithQualityWeights function (which performed in a resource and time-efficient enough manner for this smaller, one-region dataset to use as provided) rather than the customized implementation described above. Since all samples were from a single brain region, our only biological factor of interest was sex in this model:

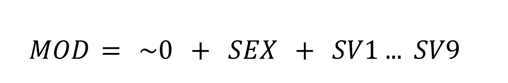

The DE table for the CMC data is available in **Supplemental Table 2.**

#### Gene Set Enrichment Analysis

In order to test for categories of biological function enriched in sex-differentially expressed genes outside of sex chromosomes, we performed Gene Set Enrichment Analysis (GSEA). All autosomal protein-coding genes with detectable brain signal, and their log2FCs, were used as input for analysis with the GSEA tool (version 4.2.3) (Mootha et al., 2003; Subramanian et al., 2005). 1000 permutations were used, and gene sets were restricted to those between 15-500 genes in size. Full list of results are available in **Supplemental Table 3**.

#### Meta-analysis

In order to get a better picture of the sex-differential biology we combined the results of the 2 independent analyses through a meta analysis. To do this we first took the intersection of the two datasets’ genes using their Ensembl ID’s, then removed all genes that didn’t all agree on the direction of logFC effect. Once we had our list of genes we combined their P-values using the *AWFisher_pvalue()* function from the *AWFisher* R package. Once the P values were calculated they were subsequently adjusted using the *p.adjust()* method from the *stats* R packages, specifically with the “BH” method, otherwise known as FDR correction. Results are available in **Supplemental Table 4.**

#### ChEA3 analysis

In order to better understand which transcription factors (TFs) may be responsible for regional sex DEGs we conducted an analysis of TF using the ChEA3 tool (Keenan et al., 2019). We first took all TFs that ranked 50 or lower using mean rank metric from the, to avoid circular logic we also excluded the GTEx coexpression heuristic from the ChEA3 tool results and recalculated the mean ranks from the other four heuristics. Some additional inclusion criteria constraints we put on the data was that TFs had to also be significantly DE in the tissue of whose DEGs were input into the ChEA3 tool, meaning that the TFs were subset only to activators. For a complete list of TFs that met these criteria see **Supplemental Table 5**.

#### Single cell enrichment analysis of genes upregulated in cortex results for each sex

The Allen Brain Atlas provides a 47,000 nucleus, single-nucleus RNA-seq dataset from 6 areas of cortex representing all major cortical cell types (Hodge et al., 2019). To identify cell types enriched for male-and female-like bulk expression patterns, we utilized the male-upregulated and female-upregulated DEG sets in combination with the scDRS tool (Zhang et al., 2022) to score single cells of adult human brain with mean-variance based control gene selection, 1500 permuted controls calculated per cell, and low-count / low-gene-total pre-filtering performed by the tool (--flag-filter-data True). Gene scores were generated from the top 1,000 autosomal genes upregulated in males OR females for cortex, for a total of 2 gene score sets. Genes are weighted by a Z-score, in this case, the Z-normalized DE significance. Subsequently, the scDRS tool’s “downstream” functions were utilized to identify genes most correlated with the weighted signatures and quantify enrichment significance and heterogeneity defined as variability in enrichment scores within pre-labeled groups, in cell types and cortical layers.

#### Candidate genes from genome-wide association study loci for autism, ADHD, and AD

Candidate genes based on proximity to genome-wide association study (GWAS) peak and transcriptome-wide association (TWAS) analysis were collected for GWAS loci associated with ADHD and autism when considered jointly (Mattheisen et al., 2022). TWAS genes with an P < 0.05 were retained for list overlap. For AD, previously identified genes/GWAS loci were collected from a recent Alzheimer’s GWAS (Bellenguez et al., 2022) specifically from supplemental table 5 (known Alzheimer’s risk genes) combined with genes from supplemental table 20(newly discovered risk genes). DEGs from each sex for each GTEx region were overlapped with the autism/ADHD and Alzheimer’s gene sets described above, using the *fisher.test()* function from the R *stats* package to calculate enrichment of sex DEGs among putative GWAS target genes.

To examine whether sex-differentially expressed Alzheimer’s genes from GTEX cortex were targets of particular mature microRNAs (miRNAs), miRDB (Chen and Wang, 2020) was used to retrieve all miRNAs predicted to regulate mRNA level for the overlapping FDR-significant genes upregulated in each sex. These predictions were then subsetted to those miRNAs retained for expression in our analysis (13 miRNAs total) and used to generate the regulatory network (**Figure 4,D**) via Cytoscape (Shannon et al., 2003).

#### Enrichment of sex DE genes for autism risk genes, postmortem case control DE genes

In order to examine how our results may relate to sex-biased disease, we compared our results with those from prior autism and Alzheimer-oriented studies. For autism rare variant genes we used SFARI Genes specifically the genescore 1 to focus on the most clearly associated genes. For autism case-control DEG sets we used two prior studies, Gandal supplementary table 3 (Gandal et al., 2022) and Werling supplemental table 2, specifically Voineagu autism up and down regulated DEGs(sheets 5 and 6)(Voineagu et al., 2011; Werling et al., 2016). For Alzheimer’s case control gene sets we used the RNAseq Harmonization Study(Sage Bionetworks, 2021), specifically the cortical region contrasts. We then subset the lists from prior studies to only contain genes that were included in our analysis, and split the lists based on direction of effect (case upregulated, control upregulated). We then tested these multiple gene sets for enrichment in each of our sex by region DEG sets using the *fisher.test()* function from the R *stats* package.

#### CPM match enrichment permutation test

To test whether the overlap between male-biased DEGs and rare causal variants in autism were not simply explained by an increase in neuronal signatures in the male samples, we tested whether random genes with similar expression in neurons would show similar enrichment to SFARI genescore 1 genes, using an approach derived from(Ouwenga and Dougherty, 2015), but updated to use single neuronal cell data to generate the random gene lists. Specifically, we utilized the Allen brain atlas cortex postmortem single-nucleus RNA data (Hodge et al., 2019) and subsetted it to only neuronal cell types. We then summed the counts, calculated the CPM for each gene, and calculated the log to obtain a vector of neuronal gene log CPMs. We then used the *density()* function to generate a probability matrix of the CPMs for each gene. We then subsetted this vector to only genes from the SFARI genescore 1 list, and used the *density()* function to generate a probability matrix of the CPM for that set of genes. Once we had both probability matrices, we sampled subsets of the neuronal genes with similar CPMs using the *sample()* function 1000 times. Each of the 1000 permuted gene sets was then tested for enrichment in the male cortex DEG set. Finally, we plotted the Odds Ratio(OR) of all 1000 permuted gene sets along with the real OR from the genuine SFARI genescore 1 genes, and calculated an empirical P value for the true set based on its placement in the random distribution.

#### Code and modules

Functions from multiple sources (edgeR, sva, parallel, rmpi, and mpich) as well as custom code were used to run this analysis. A link to a Bitbucket repository with code necessary to reproduce the reported analyses is here: https://bitbucket.org/jdlabteam/gtex_paper_analyses/src/master/.

## Results

### Processing of RNA sequencing data identifies a sex signal in most brain regions

We filtered and normalized bulk RNA-seq data from the GTEx project using a *limma* pipeline with models accounting for brain region, sex, surrogate variables (SVs), and donor effects. The full GTEXv8 release contains 2642 samples from 382 donors, spanning 13 brain regions (*n* per region ranging from 152 to 255 samples). The subset of GTEX donors included in our analyses (see *Methods*) contributed samples from seven regions on average and were mostly aged 50 to 69 years. For the *n* = 1688 samples retained from these donors, the average RNA integrity number (RIN) was 6.95 (see **Table 1** for additional demographic and sample information). We excluded all genes not meeting an inclusion threshold of 10 counts per million (CPM) in at least 19 samples. This left **14082** genes, **497** of which were allosomal. PCA clustering separates tissues primarily by region (**Figure 1A**), indicating this is the major mediator of gene expression differences across samples. We show that accounting for latent sources of variation in the data using SV adjustment does not produce outliers and largely acts to shrink the variance across samples (**Figure 1A**).

**Figure 1:**
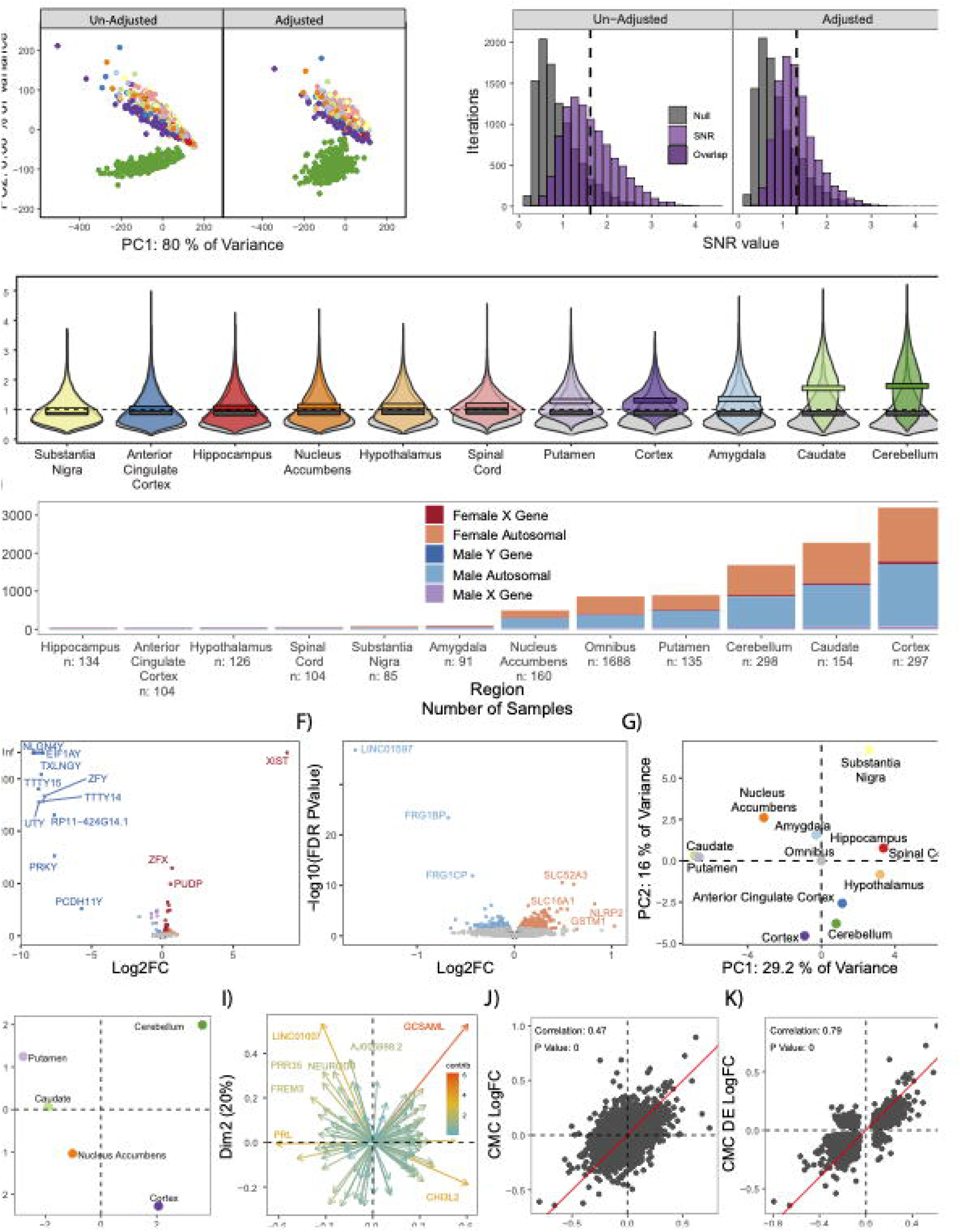
Brain regions show distinct patterns of sex-differential expression. A) PCA plot of all samples and genes before and after removing the effects of SVs. Accounting for SV’s reduces the variance in the data, and cerebellar samples (green) cluster separately from other regions *(colors as in C)*. B) SNR from 10,000 iterations, pre and post SV adjustment in cortex. SV adjustment reduces the tail of the distribution, makes the distribution more normal, and reduces mean SNR. C) SV adjusted SNR values (10,000 iterations). Each distribution is significantly different from its corresponding null distribution by Wilcox test. All regions other than substantia nigra have a mean SNR value greater than one, providing evidence that there is an expression difference between males and females in multiple brain regions. Cortex has the third highest mean SNR value and has the shortest tail, suggesting that its SNR is highly repeatable. D) Summary of DE gene count per region, including omnibus. There is an abundance of DE autosomal genes in the nucleus accumbens, cortex, cerebellum, putamen, and caudate. Sample number does not fully explain the number of DE genes in a given region. E) Volcano plot highlighting that allosomal genes follow expected trends. F) Volcano plot highlighting the autosomal genes, including noteworthy long non-coding RNA *LINC01597*. G) PCA of all LogFCs from all regions and omnibus, shows omnibus truly represents the average sex-differential expression across all brain regions. H) PCA of top 500 most variable LogFCs that were FDR significant in at least one of the “sex-differential” regions. Highlights the fact that cerebellum remains an outlier even when only considering sex differences. I) PCA plots showing the key genes that separate the SDR in the same PCA space as panel **H**. Highlights a few notable genes that can be used to distinguish regions from a sex-differential lens. J) Correlation of LogFCs between CMC, and GTEx analysis demonstrates results are robustly shared for the cortex. K) Correlation of LogFCs for DE genes between CMC and GTEx shows a high replicability in cortex.

### Sex biased signal is stronger than technical noise in the majority of tissue samples

Differential expression analysis allows for identification of genes with sex-biased expression; however, a signal to noise ratio (SNR) analysis captures pan-transcriptomic divergence between groups without using arbitrary statistical thresholds and can aid data quality assessment by quantifying signal and variance in relative terms (Lopes-Ramos et al., 2020). Moreover, this approach can be used to confirm noise-reducing effects of data pre-processing (low count removal, batch corrections, etc) (**Figure 1B**). We calculated SNR values for each region twice, once with unadjusted RNA counts and again with SV-adjusted counts. Random subsets of male and female samples were drawn 10^5^ times per region for both SV-adjusted counts and unadjusted counts (see *Methods*). We observed that adjusting for SVs makes the SNR distribution more similar to a normal distribution (**Figure 1B**). When examining the SV-adjusted counts for all regions we see that regions with simpler dissections and/or with larger total *n* (*e.g.* cerebellum) had the largest signal to noise ratios, indicating either robust sex differences or exceptionally low noise due to ease of dissection and statistical power (**Figure 1C**). The shape of the SNR distribution is also of importance as distributions approaching normal indicate that SNR values are replicable across sample subsets, while long-tailed distributions indicate that extreme findings can be driven by certain combinations of samples/donors (**Figure 1B)**. Though, unlike subsequent DE analysis, our SNR calculations did not account for random effects of donor. Substantia nigra showed a SNR of less than one, indicating that this region may not have sex-differentiated gene expression patterns, or is more difficult to dissect reproducibly and is thus noisier. For these reasons, we recommend caution in interpreting our sex DE findings from the substantia nigra. However, in remaining regions, a sex-biased signal greater than technical noise is evident, with the cortex standing out as a region with consistent and substantial sex differences.

### Sex differences in gene expression are widespread across brain regions

We next identified DE genes (FDR < .05) between males and females for each brain region, and across all regions in a general omnibus model (**Figure 1, Supplemental Table 2**). Both autosomal and allosomal genes were included in illustrations and analyses except where noted. There are DEGs (FDR < .05) both on allosomes and autosomes (**Figure 1D).** Many known X inactivation escape genes were found to be highly expressed in females, including *XIST*. As expected, DE genes shared across regions were allosomal: for example, 15% of allosomal genes (74, including all 14 chrY genes analyzed) were found to be DE in the omnibus model, consistent with base expectations of a sex DE analysis. In addition to the allosomal genes, we further identify a total of 5,182 unique autosomal genes DE in at least one of our region specific contrasts or omnibus contrast (see *Methods*: differential expression analysis), albeit at low magnitudes (mean absolute linear fold difference of autosomal DE genes was 1.158). Consistent with the region-specific nature of brain gene expression and regulation (Fullard et al., 2018; Zeisel et al., 2018; Dong et al., 2022), regional totals of DEGs were highly variable, from < 10 to thousands of autosomal DEGs. When considering all regions jointly in the omnibus model, 860 DE genes (786 autosomal) were identified, representing 6.1% of analyzed genes (5.8% of autosomal genes, **FIgure 1E, F)**. As expected, the number of DEGs is driven in part by *n*, as the correlation between sample size and number of DEGs is .64 (Spearman’s S = 101.68, P-value = .02368). The single autosomal DE gene found in all regions was the long, non-coding RNA (lncRNA) *LINC01597*, which was found to be upregulated in males. This lncRNA is relatively unannotated, but some exons are conserved across closely related species (**Supplemental Figure 2**). Surprisingly, we also found a number of chrX genes with male bias, including pseudoautosomal (shared regions of chrX and chrY) genes *PLCXD1*, *ZBED1*, and *ASMTL*, consistent with recent reports for cortex and hippocampus (but not caudate) from an independent dataset (Benjamin et al., 2022).

We further demonstrate that there are expression differences between regions, when considering raw counts, SV adjusted counts, (**Figure 1A**) or log_2_FC from differential expression (**Figure 1G,H**). Most pronounced are perhaps the difference between the cerebellum and the other brain regions. Based on the number of sex-differential genes and SNRs, we categorized the cerebellum, cortex, nucleus accumbens, putamen and caudate as sex-differential regions (SDRs) and plotted the top 100 variable log_2_FC DE genes (significant in at least one region) principle components (PC) to illustrate this grouping structure (**Figure 1H**). To examine which of these 100 most variable genes was driving the differences we plotted their relative contributions to the PCs (**Figure 1I**). One noteworthy gene: *GCSAML*, was found to only be DE (female upregulated) in the cerebellum, and very lowly expressed all other brain regions. *GCSAML* is known to be involved in the proliferation of B lymphocytes, interestingly *GCSAML* is also highly expressed in the testes and prostate of males (Fagerberg et al., 2014).

### Replication and Meta-Analysis of Cortical Findings with Commonmind Consortium

To confirm these findings were reproducible, we replicated this study using the control samples from another large (n = 278 males, 159 females) post-mortem collection (albeit limited to cortex), the Commonmind Consortium (CMC). We observe significant correlation in sex effects across these independent datasets (Figure 1J), especially when considering the union of their DEGs (Figure 1K). Using Fisher’s method to combine the P-values from both studies identifies a list of high-confidence cortical sex DEG that may be useful for future analyses **(Supplemental Table 4)**.

### Sex DE genes highlight neuronal and immune signatures in male and female brain

After establishing the detectable difference between males and females, we next investigated which biological pathways differ between the sexes. For each region we conducted Gene Ontology (GO) analysis of FDR DE genes per sex **supplemental** Figures 3-7. For cortex and omnibus models we followed up this analysis with semantic similarity clustering to reduce redundancy of terms and increase presentability (Figure 2A, full GO analyses in **supplemental** Figures 3-7). Males showed upregulation of neuronal pathways (which reproduced across both datasets) while females showed an upregulation in immune, vascular, and endothelial cell signatures, consistent with a recent report in independent brain data (Benjamin et al., 2022).

**Figure 2:**
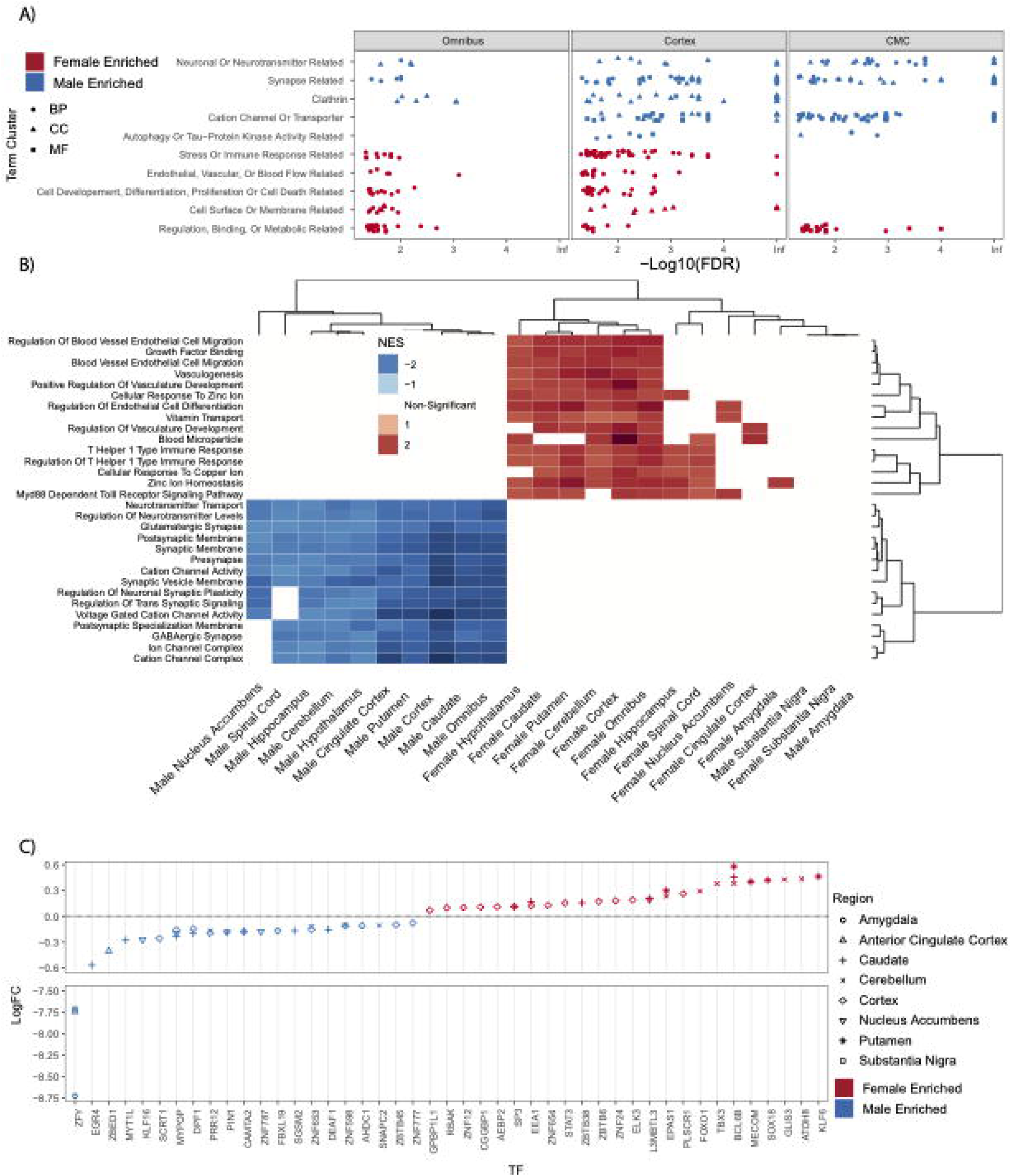
Pathways transcriptionally enriched in the male and female brain. A) Male, Female, GTEx Cortex, GTEX Omnibus and CMC Cortex up-regulated GO terms from FDR significant autosomal genes, shows male-biased and female-biased categories in cortex and omnibus. B) GSEA plot of all tissues and sexes top 15 mean highest significantly enriched categories’ NES scores from males and females clustered by sample and NES

Furthermore, although many of these ontologies were shared across regional and omnibus models (Figure 2B), some appeared to be region-specific. For example, male upregulated genes in the putamen showed mitochondrial signature and lacked the male neuronal signature found in other regions. Also, regions with low SNR values or a small number of DE genes did not reflect the general GO enrichments above. Interestingly, the cerebellum did not share the ontology findings of other regions despite having the highest regional SNR and a large number of DE genes. Indeed, essentially no terms were enriched for male-upregulated genes, and female DE genes yielded terms with only vague resemblance to the immune and endothelial signatures found in other regions. Overall, these findings provide additional evidence that indicate regional differences in sexually heterogeneous gene expression, with the cerebellum in particular being unique in its expression profile.

Next, we investigated potential drivers of the DEGs in the different regions by examining transcription factors (TFs) whose targets were significantly enriched in our DEG lists with the ChEA3 tool (Keenan et al., 2019). To identify potential regulators, we subsetted to only transcriptional activators that were themselves DE in each region. We found several high-confidence TFs, which may play a role in modulating transcription in each region (Figure 2C**,Supplemental table 5**). Notably, SRY is documented to not only be a pervasively active TF in the substantia nigra, but also plays a role in exacerbating Parkinson’s disease, which is a male-biased neurodegenerative disease (Lee et al., 2019). Likewise, the female-enriched gene, *BCL6B*, is predicted to be involved in inflammatory response, which suggests a mechanism for why many inflammatory genes are upregulated in females across regions.

### Sex specific signatures are mainly driven by differences in cell type states

Our GO results suggested many of the sex DE genes corresponded to particular cell types. We first sought to confirm this with cell type focused tools and then to interpret these results leveraging recent collections of single-cell sequencing data.

To confirm DE enrichment in the cell types suggested by ontology analysis, we intersected DE genes to mouse brain cell type markers using Cell type-Specific Expression Analysis (CSEA) (Xu et al., 2014), again revealing a neuronal signature in males and a glial/immune signature in females (**Supplemental Figure 9**). However, these initial analyses utilized tools based on a limited number of purified cell type RNA rather than true single cells (Doyle et al., 2008). Furthermore, this observation of sex DE gene enrichment in cell type-specific genes/ontology terms could be driven by either sex differences in cellular abundance or by a sex bias in cellular states within each cell type. Thus, to more rigorously identify cell types enriched for cortical sex DE genes across a larger number of cell types, and to clarify whether these enrichments represent abundance or state differences, we leveraged the Allen Brain Atlas single-cell RNA sequencing data from the adult human cortex (Hodge et al., 2019) (Figure 3A) with the scDRS tool (Zhang et al., 2022). Our aim was to identify whether or not there was an overall enrichment of sex DEGs per cortical cell type, and whether that enrichment was heterogeneous. If sex DE genes were reflective of a particular state (or subpopulation) of a cell type, enrichment heterogeneity would be expected (e.g. the sex-enriched genes would be found in cells in just a subpart of a given type), whereas homogeneous enrichment would be expected of a cell type with sex differences in cell type proportion (sex-eriched genes would be found evenly throughout all cells of a type). One example of a distinct transcriptional state / subpopulation is activated microglia, which have a different expression profile than inactive microglia (Jurga et al., 2020).

**Figure 3.**
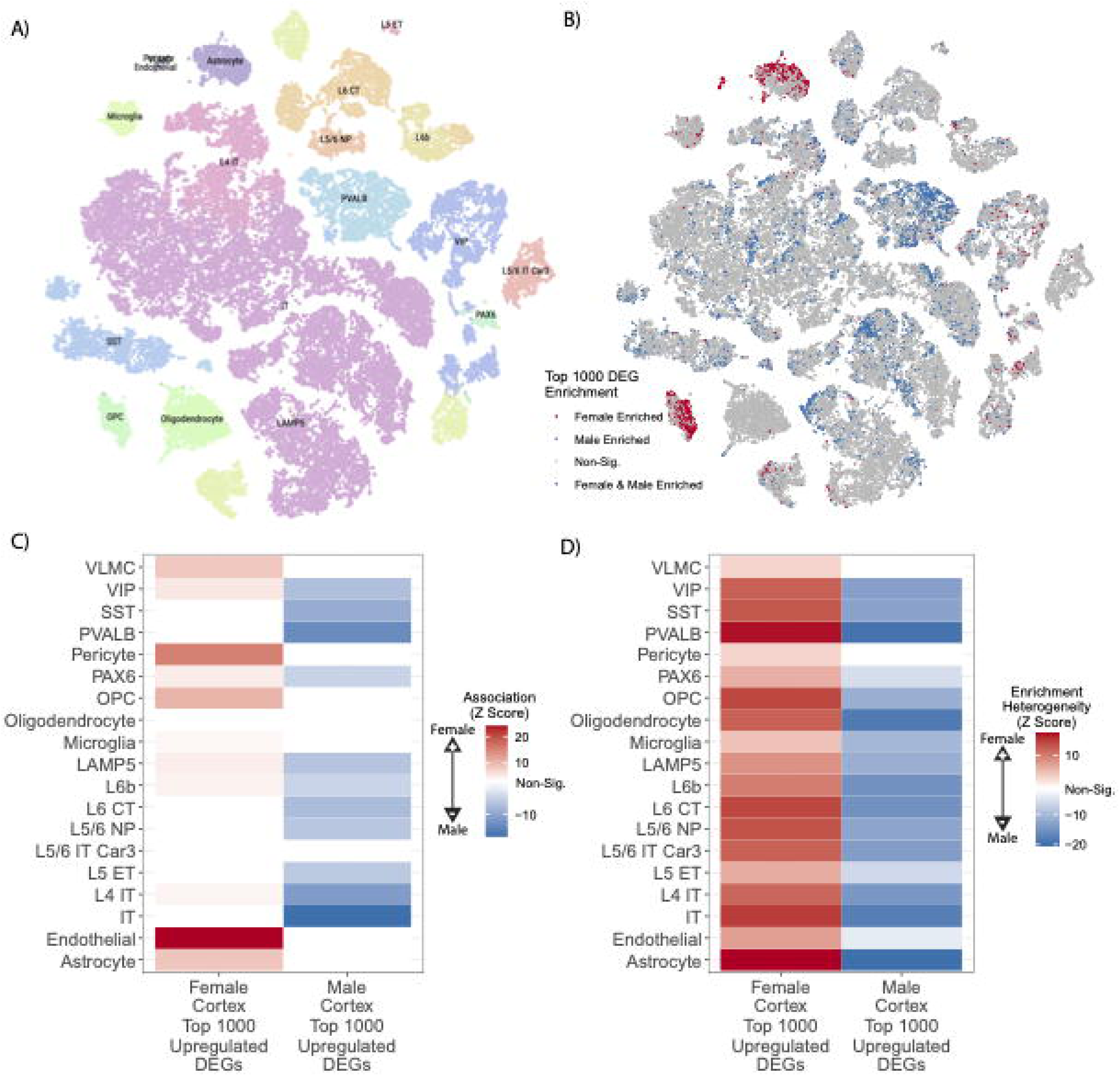
Integration with single cell data suggests sex biases in cell states. A) tSNE illustrating subclasses of single cells as identified by Hodge et al 2019; B) Male upregulated genes (blue) and female upregulated genes (red) map to individual cells in distinct clusters. C) Significance of *overall* enrichment for sex DE genes within each Allen Atlas cell subclass where white indicates non-significant enrichment after Bonferroni correction. D) Significance of *heterogeneity* in sex DE gene enrichment for each Allen Atlas subclass where white represents non-significant enrichment after Bonferroni correction.

To test this, individual cells were scored for weighted enrichment in the top 1,000 autosomal sex DE genes in cortex for each sex (Figure 3B), followed by enrichment and heterogeneity testing for each cell type using the single cell enrichment scores(Zhang et al., 2022)(Figure 3C**, 3D**). In brief, the scDRS algorithm works to test for enrichment in a cell type by generating a null distribution of randomly selected cells, each ranked with a Z-score according to their relative expression of the input set of genes; an enriched cell type has more cells that are in the upper quantiles of expression Z-scores than would be expected from the generated null distribution. Male cortex-upregulated genes were most strongly enriched in several neuronal lineages, but heterogeneously so; top *Z-*scaled male enrichments highlighted intratelencephalic neurons (13% of cells, *Z* > 18, Bonferroni-adjusted heterogeneity *p* < 0.05) and *PVALB*-expressing neurons (39% of cells, Z > 14, Bonferroni-adjusted heterogeneity *p* < 0.05) (Figure 3B**, 3C**). Female upregulated genes from cortex were most strongly enriched in 40% of single astrocytes and 63% of single oligodendrocyte progenitors (Figure 3B**, 3C**; *Z* scaled enrichment scores > 10 and 8, respectively, both Bonferroni-adjusted *p* < 0.05), each also with significant inter-cell heterogeneity (Bonferroni-adjusted *P-value*s < 0.05). These findings strongly suggest that it is particular states of most cortical neuron types, astrocytes, and oligodendrocyte precursors that drive the overall sex-differential gene expression seen in bulk RNASeq. Complete results are in **Supplemental Table 6**. To confirm these results were not spurious, we selected a random set of brain-expressed genes, arbitrarily assigned P-values from the real DEG lists and repeated both analyses. We found no significant overlap with either cell type or heterogeneity measures using this procedure.

### Lack of molecular predisposition, but increased expression of autism risk genes in male brain

We next examined how sex DE patterns relate to psychiatric diseases with sex biases in diagnosis, with two non-exclusive approaches. First, we examined prior case-control data to determine if male brains exhibit greater similarity to cases, indicating a male molecular predisposition towards an autism-like state (Werling et al., 2016; Gandal et al., 2022), while asking if female brains lean more towards an Alzheimer-like state. Secondly, we investigated whether there is any sex bias in the expression of Alzheimer-or autism-associated genes in control individuals.

#### Case-control

Female enriched genes were generally higher in autism cases than controls, contrary to a molecular predisposition hypothesis. This was true across two gene sets from autism case control studies (Figure 4A). Likewise, we found that autism downregulated cortex genes relative to controls were found to be enriched in the male upregulated cortex DEG set.

In addition to looking at gene overlap in previous bulk RNA-seq autism case control studies, we also examined the gene overlap in Alzheimer’s bulk RNA-seq case control studies. We chose an analysis (Sage Bionetworks, 2021) that used multiple cohorts of Alzheimer’s donors from different institutions and analyzed the data using several statistical models. We chose the cortical samples for each of these cohorts and used three of the analyses: diagnosis only, diagnosis by male sex only, and diagnosis by female sex only. Regardless of which cortical region examined, or which analysis used, we saw the same, clear pattern emerge. Genes upregulated in Alzheimer’s donors significantly overlap with genes upregulated in female cortex, and genes upregulated in control donors significantly overlap with male cortex (Figure 4B). This provides evidence that females have a molecular predisposition for an Alzheimer-like state.

**Figure 4.**
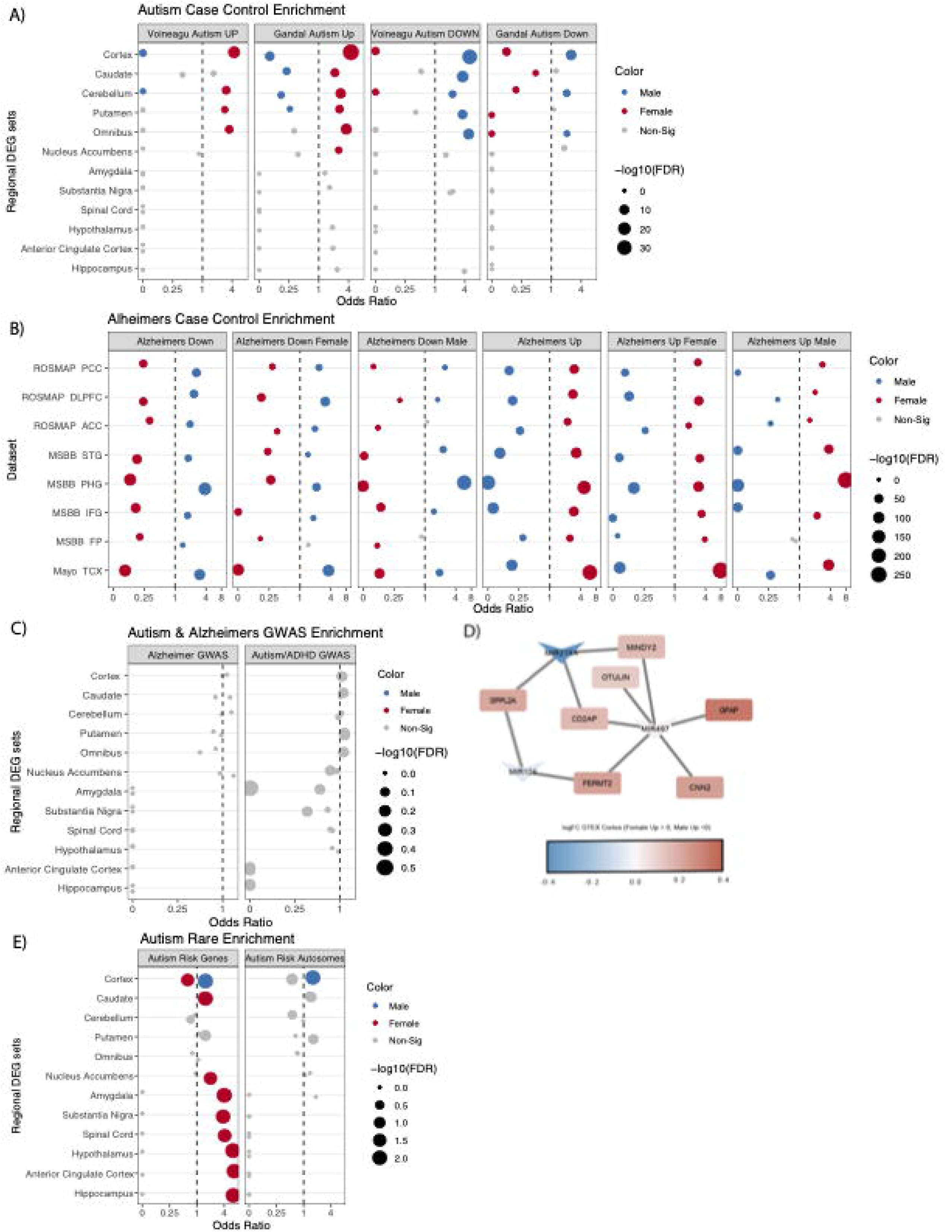
Overlap of sex differences in gene expression to postmortem case-control differences and disease gene sets. **A)** Autism significantly up-and down-regulated genes sets from case control studies. **B)** GTEx cortex male and female DEGs overlapped with male, female and sex agnostic Alzheimer’s up and downregulated genes sets from cortical samples. **C)** GWAS Alzheimer’s and autism/ADHD risk loci overlapped with male and female DEGs, shows no significant enrichment. **D)** Male DE microRNAs targets found in GWAS Alzheimer’s risk genes sets. **E)** Autism risk genes from SFARI genescore 1 both including and excluding allosomal genes, shows significant male enrichment in the cortex.

#### Common risk variants

One possible mechanism by which sex differences arise in diseases with complex (*i.e.* polygenic, common-variant mediated) heritability is through variant-mediated perturbations to baseline sex-differential gene expression patterns. For example, a set of genes expressed in a given tissue and linked to common risk variants through genome-wide association studies (GWAS) could be upregulated or downregulated as consequences of risk alleles, altering cell fate or state when collective effects surpass some threshold. Sex differences in gene expression could either buffer against or predispose toward reaching a collective effect threshold in a given pathway or cell type. For example, if disease *D* risk loci cumulatively downregulates pathway *Z*, and pathway *Z* is upregulated in healthy females in the disease tissue, then females should be resilient to genetic risk factors acting through pathway *Z*. Therefore, by looking at which common polymorphism loci show some type of nominally significant contribution to risk, we can see if any up or downregulated genes overlap and possibly identify mechanisms for risk.

We first tested whether any of the analyzed regions’ upregulated genes from each sex were enriched for genes associated with Alzheimer’s or autism by prior and recent GWAS studies (Bellenguez et al., 2022; Mattheisen et al., 2022). No significant enrichment of sex DE genes were found in autism or Alzheimer’s GWAS loci genes, however, GWAS loci each usually contain several genes, only a subset of which are related to the condition. Thus, we conducted a preliminary analysis of the subset of GWAS loci genes that did overlap with sex DEGs, to see if any patterns emerged. One interesting overlap was with a lncRNA found near the critical Alzheimer’s gene *APP–AP00023.1* – which was significantly upregulated in female GTEX omnibus and cortex. 15 additional Alzheimer’s risk genes significantly upregulated in female cortex were *RPS27L*, *SPPL2A*, *PRKD3*, *MINDY2*, *FERMT2*, *TMEM106B*, *FAM96A*, *RAB8B*, *ABCA1*, *CD2AP*, *OTULIN*, *FOXF1*, *CNN2*, *GFAP*, and *GDPD3*. To further investigate GWAS loci that did overlap with our DEGs we conducted a downstream analysis and identified enrichment for targets of the miRNA, *MIR219A2: ABCA1*, *MINDY2*, *SPPL2A*, and *CD2AP*. *MIR219A2* was upregulated in male cortex and omnibus at nominal significance (FDR .063), while the aforementioned targets were upregulated in females at corrected significance. This relationship is consistent with the repressive role of miRNAs, and suggests that *MIR219A* may confer a protective effect against Alzheimer’s by repressing these targets (Figure 4D). One study has provided evidence that MIR219A overexpression helps regulate the differentiation of oligodendrocyte precursor cells (OPCs) into oligodendrocytes, promotes remyelination, and improves cognitive function (Fan et al., 2017). Additionally, autism GWAS loci genes intersecting male-upregulated genes from GTEX cortex were enriched for putative target genes of the transcription factor and androgen receptor (AR) coregulator (Cui et al., 2011) *ZBTB7A*. *ZBTB7A* fell just short of corrected significance for upregulation in male cortex (FDR=0.06), but could hint at a similar mechanism for why this set of genes is higher in male brain.

#### Rare syndromic disorder autism genes

We also tested for enrichment of rare variant autism genes in the regional sex DEGs lists. We utilized the SFARIGene database (Abrahams et al., 2013), specifically genes with high confidence of playing a role in autism, denoted as genescore 1 (nearly all of which cause neurodevelopmental syndromes with high penetrance of both autism and Intellectual Disability). We found there to be a significant overlap of these high confidence autism risk genes across many of the regional sex DEG sets. Most commonly, the autism risk genes were enriched in females, which likely reflects the fact that many of the risk loci are found on the X chromosome (21 of the 211 autism genes were on the X chromosome), which are likewise more often upregulated in females. To remedy the expected X-linked rare variant genes driving enrichment we repeated this analysis excluding all allosomal genes (Figure 4E). We observed the male cortex was enriched for autism risk genes regardless of allosomal exclusion (Figure 4D**,E**). It should also be noted that when using the SFARIGene database list as it existed at the time of the Werling 2016 paper, we replicate the findings of Werling 2016 and find no sex DE sets to be significantly enriched. Finally, for Alzheimer’s, relatively few rare loss of function variants have been robustly associated with the disease, precluding a similar analysis.

## Discussion

This study comprehensively examines adult sex differences across brain regions and across the brain as a whole under a single unified model, providing a valuable resource for future reanalyses. It should be noted that while prior work has been done on this topic (Wapeesittipan and Joshi, 2023), none have included so many samples in a single statistical model, estimated random effects, nor compared these results specifically to Alzheimer’s and autism. Prior work has separately analyzed multiple brain regions, across multiple datasets, but as separate models, so direct comparisons across different regions are not as meaningful, and no conclusions can be drawn at the omnibus level. Interestingly, with the power of all samples available, more than 5% of genes included in this analysis showed significantly sex-biased expression, albeit often of very low magnitudes in the omnibus model. Interpretation of these results are complex and can vary greatly depending on selected log_2_FC thresholds. Thus, care must be taken in selecting significance and log_2_FC thresholds most relevant to a given line of inquiry or quantitative approaches leveraging all available log_2_FC values should be taken. In addition to the GTEx omnibus model we also present the individual analyses for each brain region (**Supplemental Tables 1**).

Looking across regions, *LINC01597* is a newly identified sex DE gene of particular interest, as it shows similar male-increased expression patterns across all models to that of an allosomal gene despite not having homology to any known Y region (Figure 1F). This extreme sex bias can be seen in other large human genetic studies (García-Pérez et al., 2023; Reynolds and Niedbalski, 2023). This could be a novel example of a uniquely regulated pseudoallosomal gene that may have important function, considering its high relative expression in the brain (Fagerberg et al., 2014) and pituitary (Carithers et al., 2015). The *LINC01597* gene is found near the centromere of chromosome 20 and shows some conservation (**Supplemental** Figure 2). Search of *LINC01597* sequence to telomere to telomere (T2T) genome using UCSC BLAT search confirmed no homology to Y genes, ruling out mis-mapping of Y-derived reads as driving this finding. Considering that some lncRNAs serve as important mediators of sex specific gene expression, such as *XIST*, and have been associated with increased risk of depression and cancer (Żylicz et al., 2019; Issler et al., 2020; Rubin et al., 2020), *LINC01597* may merit additional investigation.

From pathway/ontologies analyses, our findings indicate a significant upregulation of genes associated with the Homotypic Fusion and Protein Sorting (HOPS) complex within the male cortex as well as tau protein kinase activity (Figure 2A). The HOPS complex is recognized for its involvement in autophagosomal activity, a process crucial for removing debris from the cell. Dysregulation of the HOPS complex can potentially lead to neuronal cell death. Notably, if females exhibit a relatively lower HOPS complex expression or a reduction in the amount of tau-kinase activity, these features may provide partial explanations for the higher prevalence of Alzheimer’s disease in the female population.

Examination of this data with regards to cell types suggests states of particular cell types may also contribute. We observe evidence of sex-differential cell states in neurons, with males having an upregulation for certain neurotransmitters and synapse formation genes while females show evidence for possessing distinctive vascular, endothelial, and immune signatures. Specifically, our findings contribute evidence to a growing body of knowledge that indicates the female brain has increased levels of immune activity, either via an increased abundance of immune cells (particularly microglia) or via a more active state of these cells (Trabzuni et al., 2013; Hanamsagar et al., 2017; McCarthy et al., 2017; Nelson and Lenz, 2017). Our subsequent analyses incorporating single cell data suggest that females do not have an increased abundance of microglia cells; rather that these cells are in a different state (Figure 3C**,D**). Distinct microglia states are strongly implicated in Alzheimer’s risk and progression through both human genetics and pathology (Paranjpe et al., 2021; Coales et al., 2022). For example, high-throughput analysis of microglial morphometrics in mice also indicated female microglia are in a more disease-like state, and more rapidly shift into this state in progress of disease models as well (Colombo et al., 2022). If microglia in the human female brain are also already slightly shifted toward such states, this could enhance the risk of developing disease. This shift would have implications for diseases and disorders beyond Alzheimer’s disease, including Multiple Sclerosis (Fish, 2008).

There may be an opportunity to consider treatments or risk mitigation approaches that are informed by the sex of the person, although we are unaware of exactly how these sex differences in gene expression are being driven. One possibility is miRNA 219A2, which we observed as being possibly sex-biased. If so, this could be male-protective via degradation of Alzheimer’s implicated transcripts, providing one possible explanation of increased female risk for Alzheimer’s disease. However, other possible mechanisms may include specific TFs, like *BCL6B, SCRT1* highlighted here, or others in the cacophony of regulators that are downstream of the allosomes, via sex hormones, or a nebulous combination of environmental and sociological factors, or a nuanced and complex mixture of all of the above. Thus, the female gene-expression bias towards an Alzheimer-like state might suggest any molecular predisposition is acting through multiple convergent molecular pathways.

Notably, elderly individuals may exhibit an augmented immune response in brain tissue, and since females tend to live longer, it is plausible that the observed immune signature may be caused by data skewed toward older donors. However, to rule out this possibility, we reran the analysis with age as a covariate and the findings remained. Thus, at least within the age ranges present here, this seemed to be a *bona fide* sex difference rather than age effect.

Somewhat more difficult to interpret is the evidence for a change in neuron and neurological factors seemingly upregulated in males. Our single cell data analysis seems to suggest this sex bias is due to a change in state (Figure 3). One hypothesis could be that neurons in males brains are regulated by a different proteasomal and hormonal environment, which causes these neuronal pathways to appear differentially expressed, perhaps including the HOPS pathway discussed above.

Unlike our Alzheimer’s analysis, our case-control gene expression findings did not support a male molecular predisposition as driving the sex difference in autism prevalence, in contrast to prior work (Kissel and Werling, 2020). While the sex DE here was well powered, our data sets sampled an older population, thus differences in either power or age of samples may explain the difference in findings from the prior study. Moreover, the female component of our dataset is largely postmenopausal in age, and thus would miss gene expression effects of many circulating hormones. Thus, future well-powered studies should test whether this same sex DE pattern holds in younger brains, especially those from ages when autism is diagnosed.

It was interesting that outside of the X chromosome, rare-variant causal genes for autism overlap with male-biased sex-differential genes. This could potentially simply reflect the higher male expression of neuronal genes, combined with the known bias of autism genes towards neuronal expression. To test this, we generated 1000 random gene sets of the same number and expression level in single cell neuronal data, as the autism gene list and ran enrichment testing in the male cortex DEGs. We did find these random neuron-biased lists were slightly enriched in male brain DEGs (mean odds ratio of ∼1.2), yet they rarely matched the odds ratio observed in the true autism gene list (**Supplemental** figure 10). In light of this, we could interpret this overlap as a greater dependence on each of these genes in the male cortex, though of course any individual carrying these mutations is only losing one of these genes, not the whole set. Postmortem studies of individuals with these rare mutations may be informative in better understanding this result.

Overall this work provides a robust analysis of adult human RNA expression across multiple brain regions as a resource for future use. Furthermore, these findings highlight both individual genes and specific pathways to consider when trying to better understand sex biases in health and disease.

## Data Availability

No data were produced for this study - it was a reanlaysis of existing data sources.

## Acknowledgements

We’d like to thank Tychelle Turner, Yating Liu, Oscar Hariri, Jeffrey Leek, Andrew Jaffe, Eugenia Radulescu, Christopher Maher, Sarah Koester, the members of the Gateway Working Group on Sex Differences in Autism, and Dan Geschwind for discussions and advice. We’d also like to thank Brian Koebbe, Eric Martin, and Kyle Kniepkamp for technical support. This work was funded by the Simons Foundation (734069). The Genotype-Tissue Expression (GTEx) Project was supported by the Common Fund of the Office of the Director of the National Institutes of Health, and by NCI, NHGRI, NHLBI, NIDA, NIMH, and NINDS. Some data used for the analyses described in this manuscript were obtained from dbGaP accession number <u>phs000424.v9.p2</u> on **12/13/2021.**

The results published here are also in part based on data obtained from the AD Knowledge Portal (https://adknowledgeportal.org). Data generation was supported by the following NIH grants: P30AG10161, P30AG72975, R01AG15819, R01AG17917, R01AG036836, U01AG46152, U01AG61356, U01AG046139, P50 AG016574, R01 AG032990, U01AG046139, R01AG018023, U01AG006576, U01AG006786, R01AG025711, R01AG017216, R01AG003949, R01NS080820, U24NS072026, P30AG19610, U01AG046170, RF1AG057440, and U24AG061340, and the Cure PSP, Mayo and Michael J Fox foundations, Arizona Department of Health Services and the Arizona Biomedical Research Commission. We thank the participants of the Religious Order Study and Memory and Aging projects for the generous donation, the Sun Health Research Institute Brain and Body Donation Program, the Mayo Clinic Brain Bank, and the Mount Sinai/JJ Peters VA Medical Center NIH Brain and Tissue Repository. Data and analysis contributing investigators include Nilüfer Ertekin-Taner, Steven Younkin (Mayo Clinic, Jacksonville, FL), Todd Golde (University of Florida), Nathan Price (Institute for Systems Biology), David Bennett, Christopher Gaiteri (Rush University), Philip De Jager (Columbia University), Bin Zhang, Eric Schadt, Michelle Ehrlich, Vahram Haroutunian, Sam Gandy (Icahn School of Medicine at Mount Sinai), Koichi Iijima (National Center for Geriatrics and Gerontology, Japan), Scott Noggle (New York Stem Cell Foundation), Lara Mangravite (Sage Bionetworks).

## Supplemental Tables

https://wustl.app.box.com/folder/212289113830

**Supplemental Table 1. Table of complete results of GTEx DE analysis.** For each contrast included in the analysis (both region and omnibus) each genes logFC, P-value, and gene related metadata.

**Supplemental Table 2. Table of complete results of CMC DE analysis.** For each gene included in the analysis, logFC, P-value, average expression and gene related metadata.

**Supplemental Table 3. Table of complete results of GSEA analysis of GTEx data.** For each sex, for each region, all the enriched gene set categories along with their NES scores, P-values, gene set size and other key information.

**Supplemental Table 4. Complete results of meta analysis, combining P-values from GTEx DE analysis and CMC DE analysis for Cortex.** For each gene included in both CMC and GTEx analyses that agree on direction of effect, combined P-value, and gene associated meta are available.

**Supplemental Table 5. Results from ChEA3 analysis.** For each regional DEG set a list of the most enriched activators, along with their mean rank, and gene information pulled from the DE analysis.

**Supplemental Table 6. Complete results from scDRS analysis.** Complete scDRS enrichment results for Allen brain atlas cortex single-nucleus data, including male and female enrichments for cell sub-class and cortical layers.

## Supplemental Figures

**Supplemental Figure 1.**
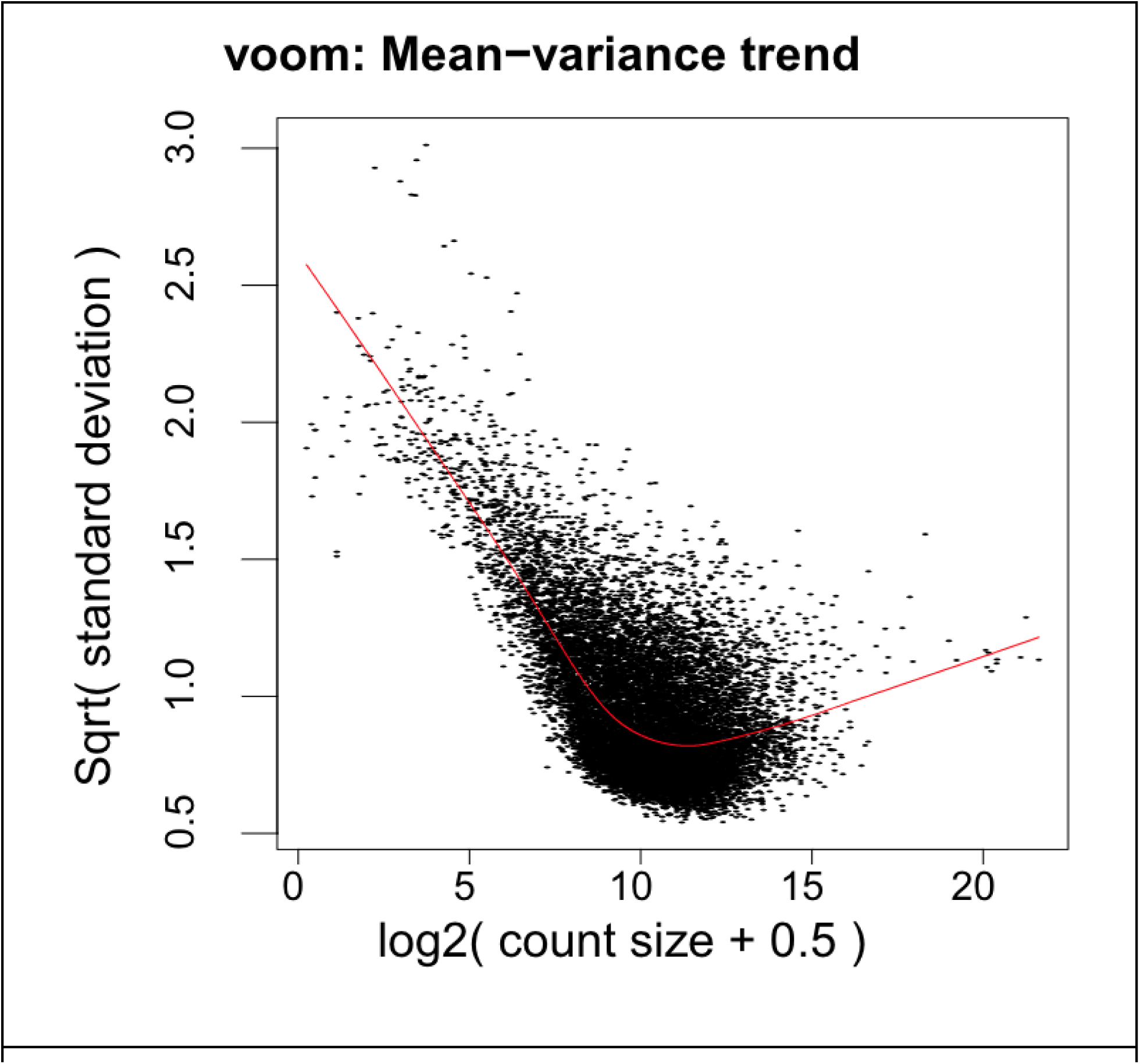
Mean variance trend. GTEx genes that passed filtering steps and their mean variance trend; shows most genes have a squared standard deviation of .5 to 1.5.

**Supplemental Figure 2:**
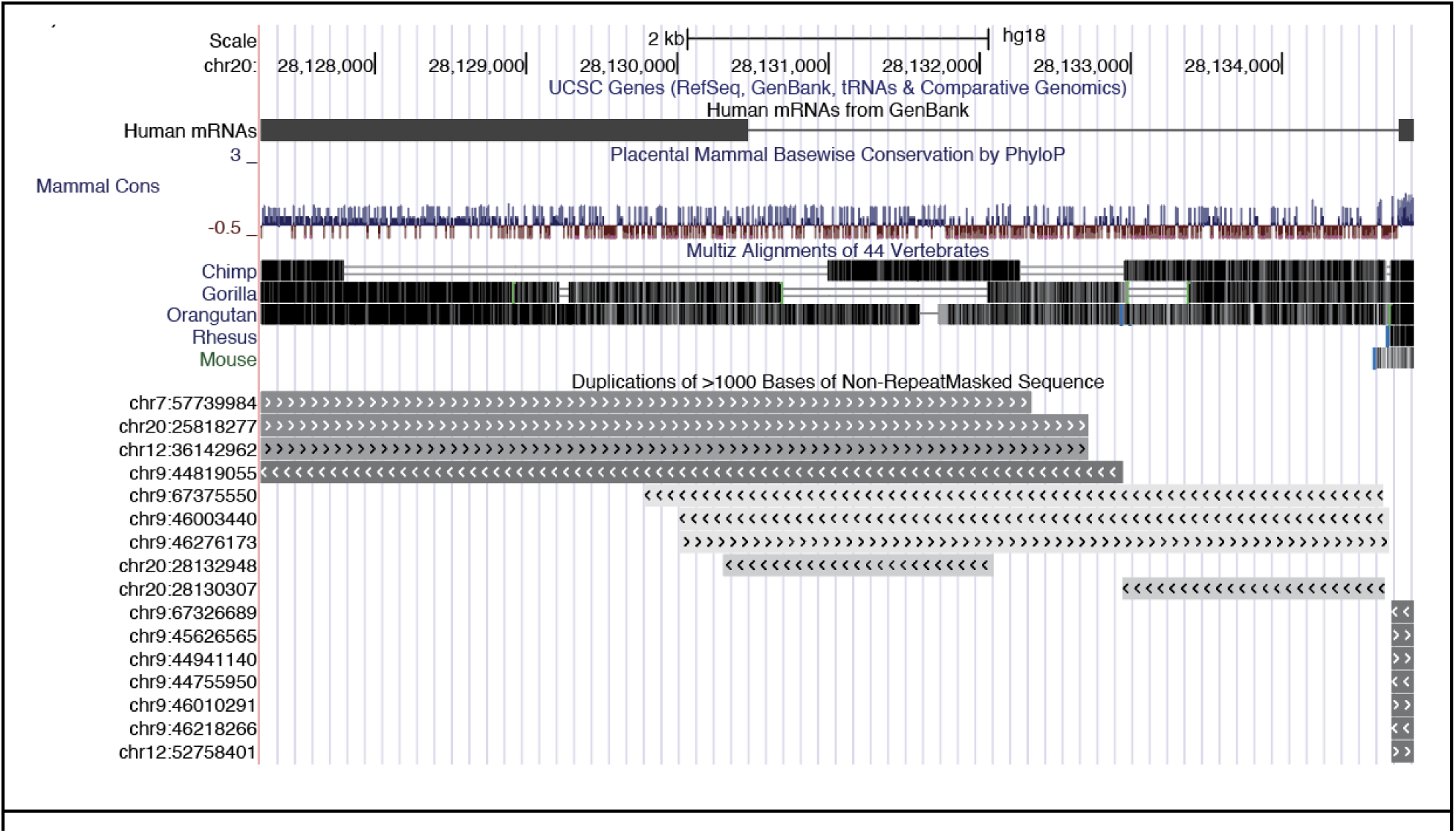
Conservation of LINC01597. UCSC genome browser track of *LINC01597.* Phylop and alignment tracks show conservation of exons. Segmental duplication track shows there are duplications of some of this region on other autosomes, but no duplication mapping to the Y chromosome. Adapted from UCSC browser (Kent et al., 2002).

**Supplemental Figure 3:**
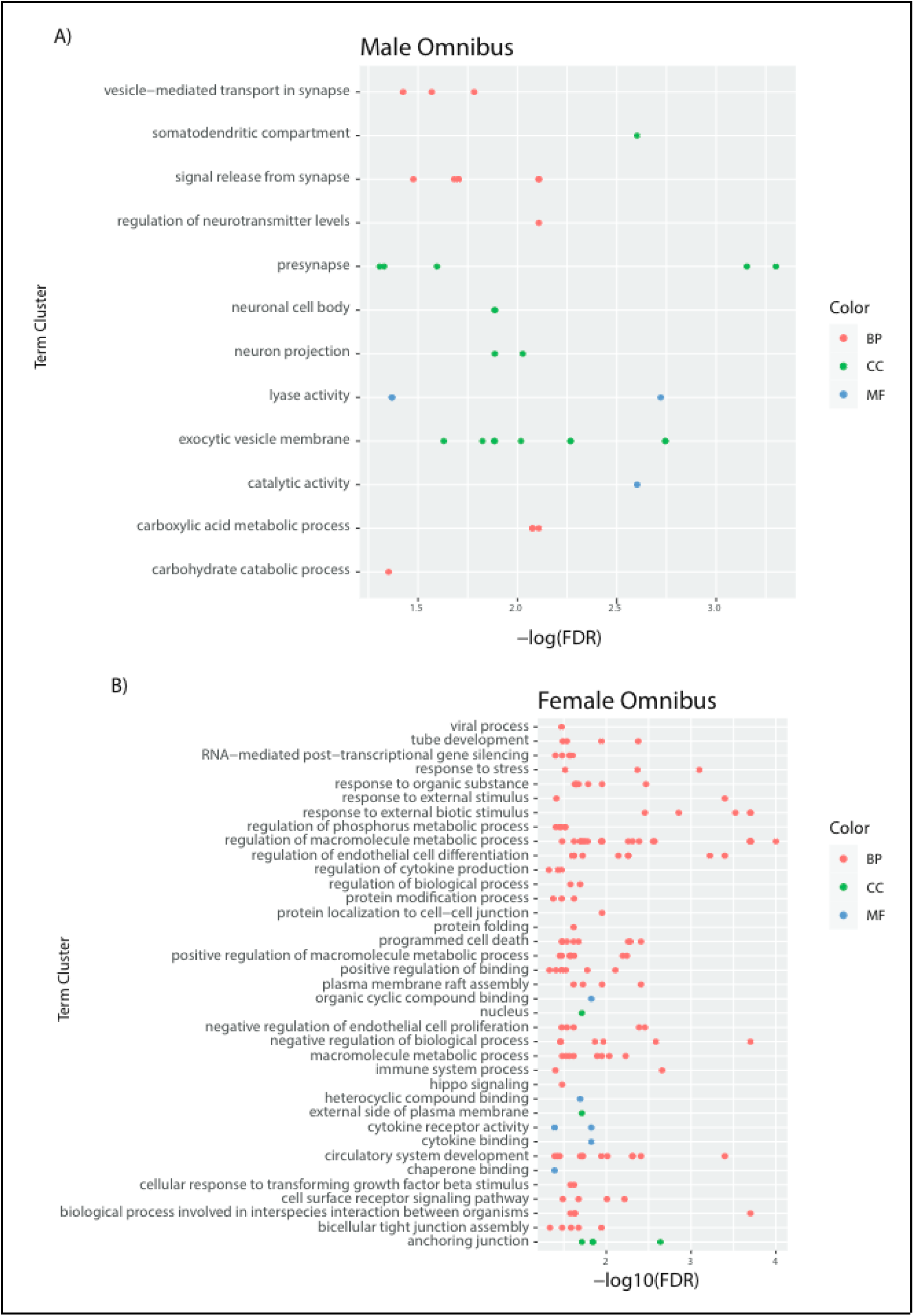
Gene Ontology plot of Omnibus results. **A)** Male Omnibus DEG autosomal genes significant (FDR < .05) GO enrichment term clusters and FDR value **B)** Female Omnibus DEG autosomal genes significant (FDR < .05) GO enrichment term clusters and FDR value

**Supplemental Figure 4:**
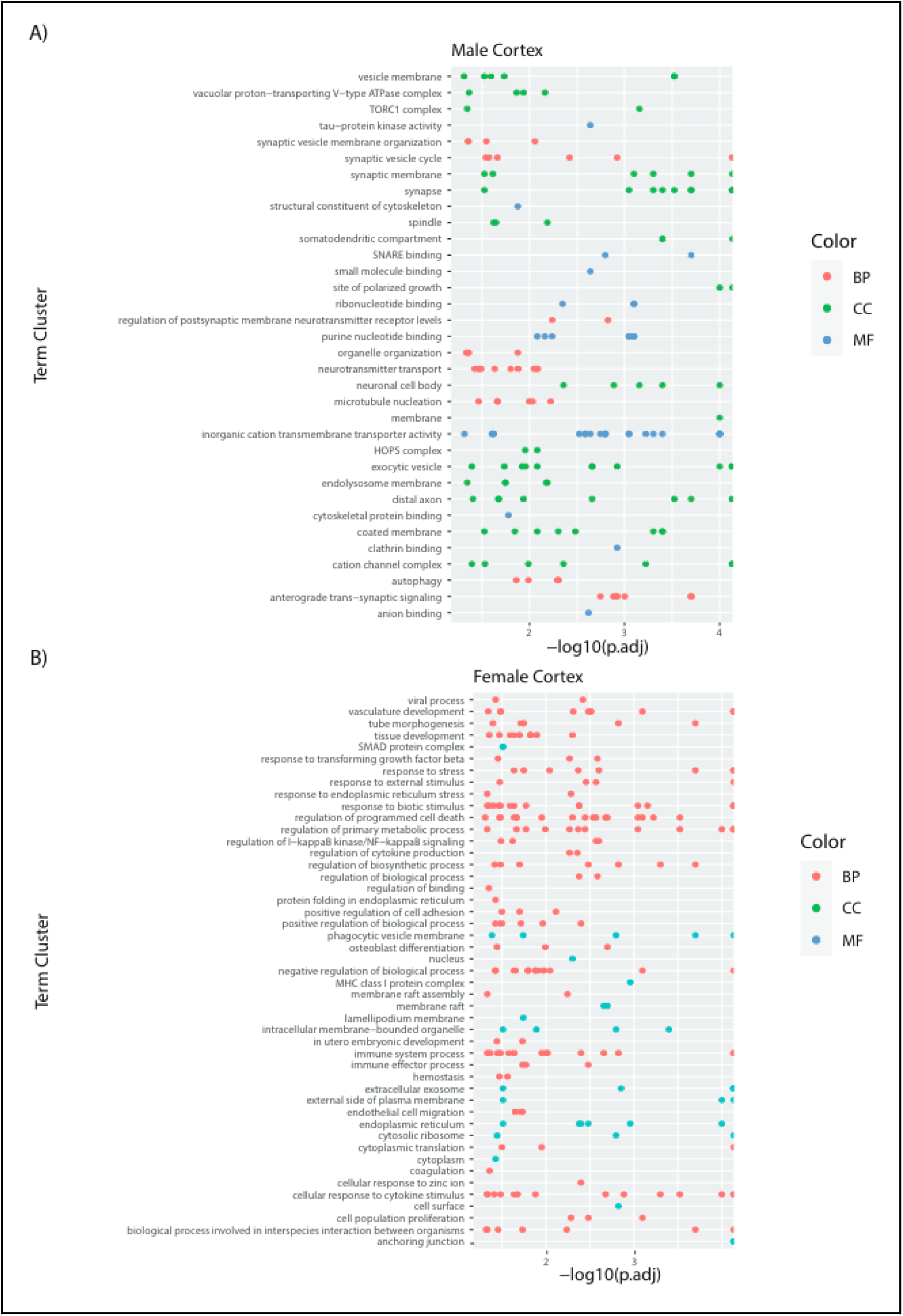
Gene Ontology plot of Cortex results. **A)** Male Cortex DEG autosomal genes significant (FDR < .05) GO enrichment term clusters and FDR value **B)** Female Cortex DEG autosomal genes significant (FDR < .05) GO enrichment term clusters and FDR value

**Supplemental Figure 5:**
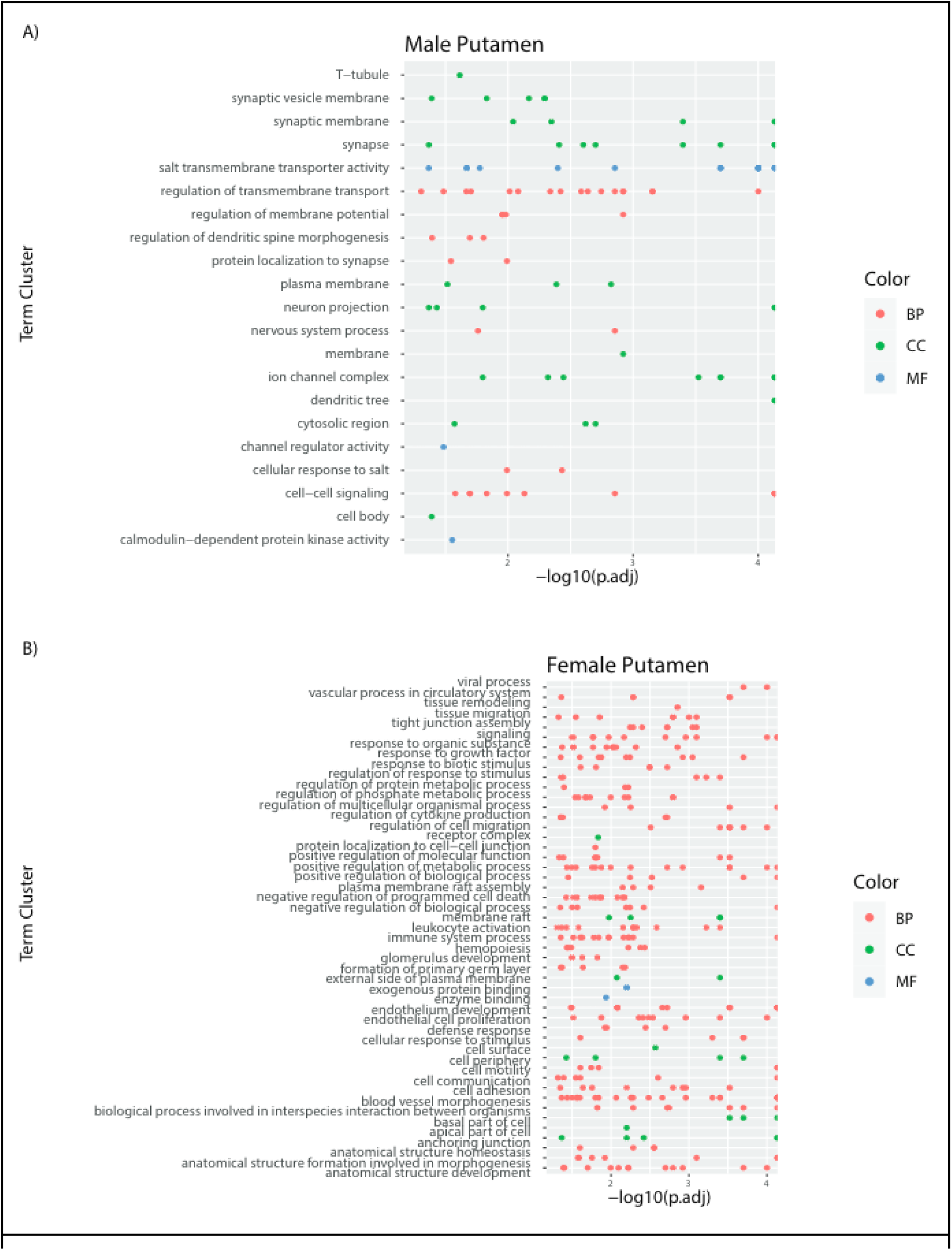
Gene Ontology plot of Putamen results. **A)** Male Putamen DEG autosomal genes significant (FDR < .05) GO enrichment term clusters and FDR value **B)** Female Putamen DEG autosomal genes significant (FDR < .05) GO enrichment term clusters and FDR value

**Supplemental Figure 6:**
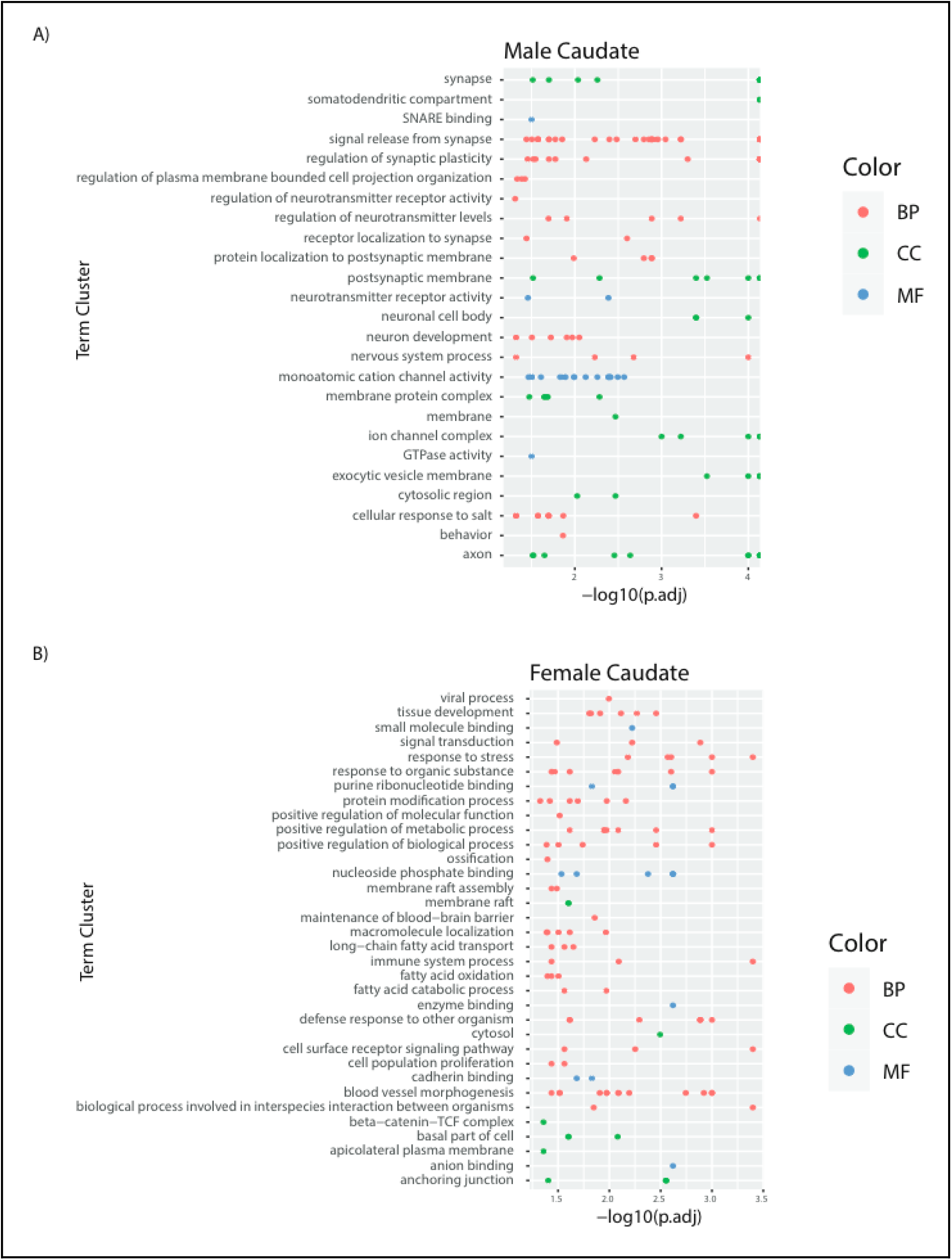
Gene Ontology plot of Caudate results. **A)** Male Caudate DEG autosomal genes significant (FDR < .05) GO enrichment term clusters and FDR value **B)** Female Caudate DEG autosomal genes significant (FDR < .05) GO enrichment term clusters and FDR value

**Supplemental Figure 7:**
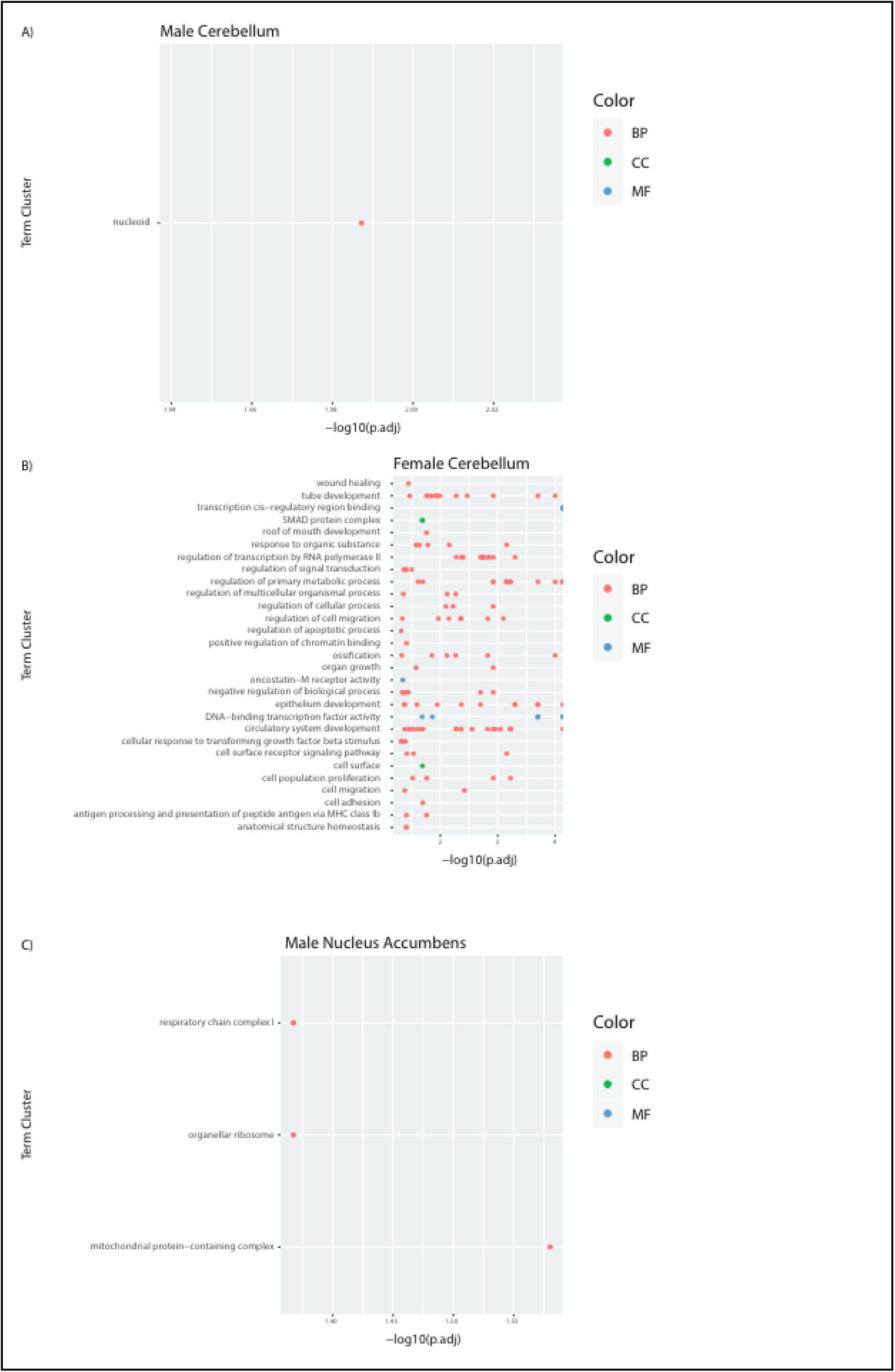
Gene Ontology plot of Cerebellum and Nucleus **Accumbens results**. **A)** Male Cerebellum DEG autosomal genes significant (FDR < .05) GO enrichment term clusters and FDR value **B)** Female Cerebellum DEG autosomal genes significant (FDR < .05) GO enrichment term clusters and FDR value **C)** Male Nucleus Accumbens DEG autosomal genes significant (FDR < .05) GO enrichment term clusters and FDR value

**Supplemental Figure 8:**
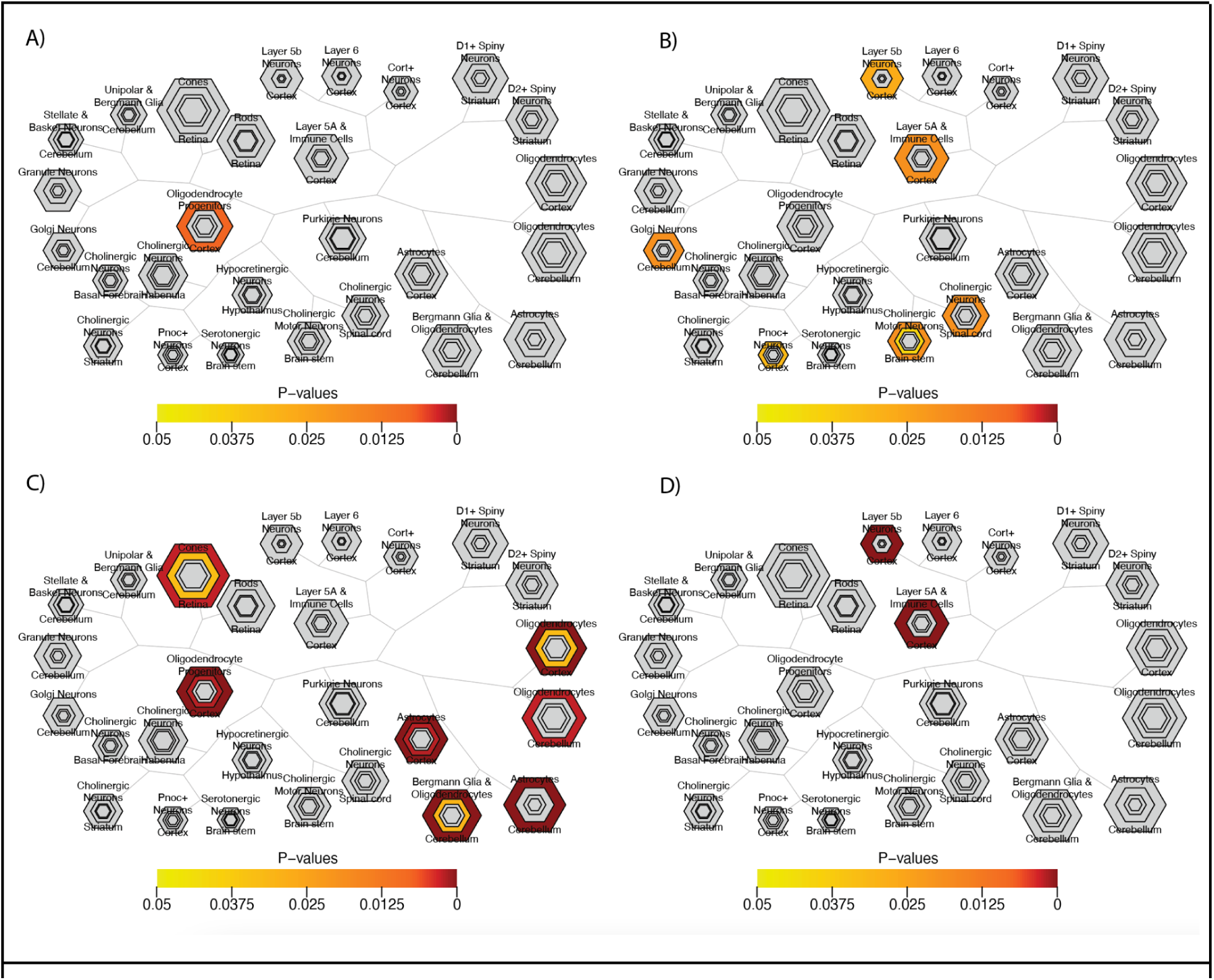
Cell type-Specific Expression Analysis suggests enriched glial signature in female cortex and neuronal signature in male cortex. a) Female omnibus protein coding genes using CSEA tool at a FDR .05 threshold, shows weak enrichment of OPCs. b) Male omnibus protein coding genes using CSEA tool at a FDR .05 threshold, shows enrichment for several classes of neurons. c) Female cortex protein coding genes using CSEA tool at a FDR .025 threshold, shows enrichment for several classes of brain immune cell types, as well as strong enrichment for OPCs. d) Male cortex protein coding genes using CSEA tool at a FDR .025 threshold, shows enrichment for layer 5b and 5a neuron subtypes. Figures generated by CSEA too(Xu et al., 2014).

**Supplemental Figure 9:**
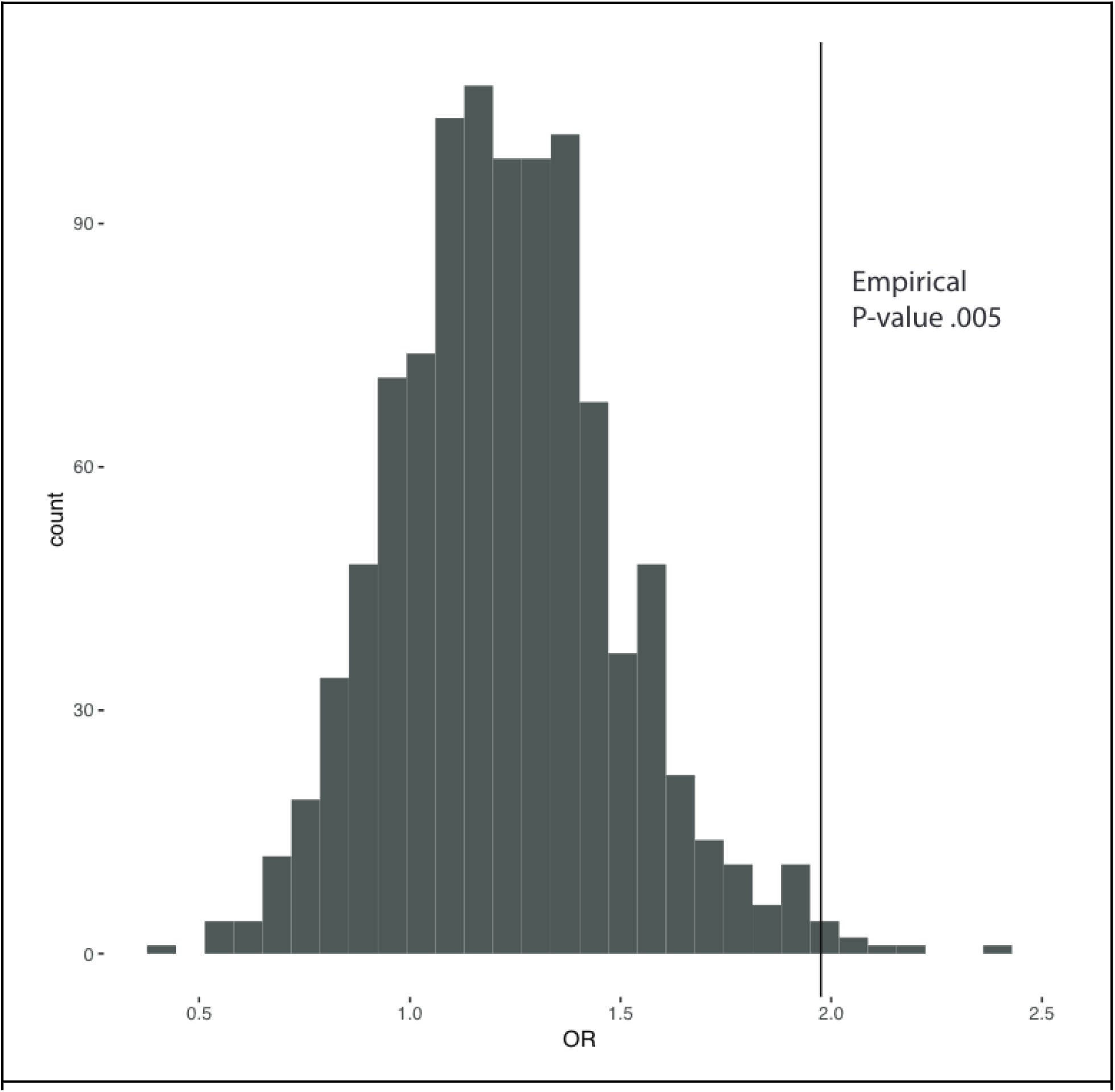
Odds ratio distribution of SFARI CPM matched neuron expressed genes. OR distribution of enrichment of random genesets with a similar CPM distribution to SFARI genescore 1 genes (vertical line) in postmortem cortex neuron data from the Allen Brain Atlas (Hodge et al., 2019). Shows that enrichment of rare variant genes in male cortex is not due to the neuronal biased male signature alone.

